# COVID-19 and pregnancy: An umbrella review of clinical presentation, vertical transmission, and maternal and perinatal outcomes

**DOI:** 10.1101/2021.04.29.21256327

**Authors:** Agustín Ciapponi, Ariel Bardach, Daniel Comandé, Mabel Berrueta, Fernando J. Argento, Federico Rodriguez Cairoli, Natalia Zamora, Victoria Santa María, Xu Xiong, Sabra Zaraa, Agustina Mazzoni, Pierre Buekens

**Affiliations:** Instituto de Efectividad Clínica y Sanitaria (IECS-CONICET), Buenos Aires, Argentina; School of Public Health and Tropical Medicine, Tulane University, New Orleans, USA; School of Pharmacy, University of Washington, Seattle, USA

**Keywords:** Coronavirus, Coronavirus Infections, COVID-19, Diagnostic Techniques, Obstetrical and Gynecological, Disease Susceptibility, Female, Humans, Infant Health, Infant, Newborn, Infectious Disease Transmission, Vertical, Maternal Health, Mortality, Pregnancy, Pregnancy Complications, Pregnancy Complications, Infectious, Pregnancy Outcome, Pregnant Women, Prevalence, Primary Prevention, Prognosis, Risk, Risk Factors, SARS-CoV-2, Signs and Symptoms, Treatment

## Abstract

**Background:** We conducted an overview of systematic reviews (SRs) summarizing the best evidence regarding the effect of COVID-19 on maternal and child health following Cochrane methods and PRISMA statement for reporting (PROSPERO-CRD42020208783).

**Methods:** We searched literature databases and COVID-19 research websites from January to October 2020. We selected relevant SRs reporting adequate search strategy, data synthesis, risk of bias assessment, and/or individual description of included studies describing COVID-19 and pregnancy outcomes. Pair of reviewers independently selected studies through COVIDENCE web-software, performed the data extraction, and assessed its quality through the AMSTAR-2 tool. Discrepancies were resolved by consensus. Each SR’s results were synthesized and for the most recent, relevant, comprehensive, and with the highest quality, by predefined criteria, we presented GRADE evidence tables.

**Results:** We included 66 SRs of observational studies out of 608 references retrieved and most (61/66) had “critically low” overall quality. We found a relatively low degree of primary study overlap across SRs. The most frequent COVID-19 clinical findings during pregnancy were fever (28-100%), mild respiratory symptoms (20-79%), raised C-reactive protein (28-96%), lymphopenia (34-80%), and pneumonia signs in diagnostic imaging (7-99%). The most frequent maternal outcomes were C-section (23-96%) and preterm delivery (14-64%). Most of their babies were asymptomatic (16-93%) or presented fever (0-50%), low birth weight (5-43%) or preterm delivery (2-69%). The odds ratio (OR) of receiving invasive ventilation for COVID-19 versus non-COVID-19 pregnant women was 1.88 (95% Confidence Interval [CI] 1.36-2.60) and the OR that their babies were admitted to neonatal intensive care unit was 3.13 (95%CI 2.05-4.78). The risk of congenital transmission or via breast milk was estimated to be low, but close contacts may carry risks.

**Conclusion:** This comprehensive overview supports that pregnant women with COVID-19 may be at increased risk of adverse pregnancy and birth outcomes and low risk of congenital transmission.

**Tweetable abstract:** A new comprehensive overview of the best available systematic reviews about the harmful effects of COVID-19 on maternal/perinatal outcomes.

## Introduction

Women undergoing pregnancy, and those at the time of childbirth and puerperium constitute potentially vulnerable populations for COVID-19. Although our understanding of this disease is growing every day, many answers are still needed about the diagnostics and the clinical management methods in these groups, the impact of the disease in pregnant women and newborns, and the potential of mother-to-child transmission.

Although some living guidelines on COVID-19 target the pregnancy population, several clinical questions regarding pregnancy and childbirth remain unanswered [1]. The rate of covid-19 in pregnant and recently pregnant women attending or admitted to hospital for any reason was around 10%. Pregnancy, in general, does not significantly increase the risk of being infected by SARS-CoV-2 [2].

The World Health Organization (WHO) stated that pregnant women or recently pregnant women who are older, overweight, and have pre-existing medical conditions such as hypertension and diabetes seem to have an increased risk of developing severe COVID-19[3]. In general, there is a consensus that breastfeeding should be promoted due to its mutual benefits. However, it is not well known whether the virus can be transmitted through breastmilk [4].

Systematic reviews (SRs) constitute an organized effort to collect and comprehensively synthesize the best available evidence on a given topic. Through this panoramic review of SRs, we aimed to answer a series of clinical questions about COVID-19 and pregnancy by summarizing the body of evidence and highlighting the best reviews in completeness and methodological quality.

## Objectives

To summarize the clinical presentation, vertical transmission, and maternal and perinatal outcomes in pregnant women with COVID-19 and their neonates.

## Methods

We performed an overview of SRs or umbrella review (PROSPERO Registration number CRD42020208783) following Cochrane methods[5] and the Preferred Reporting Items for systematic Reviews and Meta-Analyses (PRISMA) statement[6] and a specific guideline for overviews[7] **(S1. PRISMA for Overviews of systematic reviews checklist)** for reporting.

### Eligibility criteria

We included SRs that met the Database of Abstracts of Reviews of Effects (DARE) criteria[8]: 1) reported eligibility criteria, 2) adequate search, 3) data synthesis, 4) risk of bias assessment and/or 5) individual description of included studies.

To be included, SRs had to meet at least four of these criteria, the first three of which were mandatory. The exposures of interest were defined as diagnosis of SARS-CoV-2 infection, SARS-CoV-2 risk factors, diagnostic tests, or treatments. Pregnant women without interventions or exposures under study, including active or inactive comparators, usual care, or placebo, were defined as comparison groups. Any pregnancy or neonatal outcomes, including clinical presentation, laboratory, and radiological findings, were included **(S2. Outcomes of interest)**.

### Search strategy

From January to October 2020 an experienced librarian searched the Cochrane Library, MEDLINE, EMBASE, Latin American and Caribbean Health Sciences Literature (LILACS), Science Citation Index Expanded (SCI-EXPANDED), China Network Knowledge Information (CNKI), WHO Database of publications on SARS-CoV-2, EPPI-Centre map of the current evidence on COVID-19, guidelines published by national and international professional societies (e.g., ACOG, RCOG, FIGO), pre-print servers (ArXiv, BiorXiv, medRxiv, search.bioPreprint), and COVID-19 research websites (PregCOV-19LSR, Maternal and Child Health, Nutrition: John Hopkins Centre for Humanitarian health, the LOVE database) We also searched the reference lists of included SRs. No language or publication status restrictions were applied (The whole search strategy is presented in the **S3. Search strategy**).

### Study selection data extraction and quality appraisal

Pairs of reviewers independently screened titles and abstracts. We retrieved all potentially relevant full-text study reports/publications, and two reviewers independently evaluated the full-texts, recording the reasons for exclusion of the ineligible studies. Disagreements were resolved through discussion of the review team. This process was performed using the web-based software COVIDENCE [9].

Pairs of reviewers independently performed the data extraction through an online extraction form previously piloted in five studies. We recorded publication date, number of included studies, number of included participants, quality items, and the components of our research questions (population, exposition, comparisons, and outcomes). Discrepancies were resolved by consensus.

Pairs of reviewers independently assessed the quality of SRs through the AMSTAR-2 tool [10]. The instrument has 16 items. It is not intended to generate an overall score but provides a categorical rating based on critical domains: protocol register, adequacy of the literature search, justification for excluding individual studies, risk of bias from individual studies being included, appropriateness of meta-analytical methods, consideration of risk of bias when interpreting the results, assessment of publication bias. The overall quality or confidence in the results of the review can be rated as “high” (no or one non-critical weakness), “moderate” (more than one non-critical weakness), “low” (one critical flaw with or without non-critical weaknesses), and “critically low” (more than one critical flaw with or without non-critical weaknesses). Discrepancies were resolved by consensus. We did not assess the quality of the included primary studies in the SRs nor the quality of reporting of each SR.

### Synthesis of results

An analysis of the overlap of the primary studies included by each systematic review was performed. Only primary articles with DOI numbers were included for this analysis. We presented all the outcome measures reported in the SRs with proportions, relative risks, odds ratios, risk difference, and/or ‘number needed to treat’ mean differences, standardized mean differences, with 95% confidence intervals.

The purpose of our study is to present and describe the current body of SRs evidence on COVID-19 in maternal and neonatal health. Therefore, we synthesized the results of all relevant SRs, regardless of topic overlap, considering that re-extracting and re-analyzing outcome data from non-overlapping studies was unfeasible and outside the scope of this overview. Additionally, for this scoping synthesis, we selected the best SR that answers a specific question according to pre-defined prioritizing criteria: most relevant, most comprehensive, most recent, and highest quality determined by AMSTAR-2 [10]. For these prioritized reviews, pairs of reviewers independently assessed the risk of bias of the priori SR using the tool Risk of Bias in Systematic Review (ROBIS) [11] and the GRADE approach evaluating the certainty of evidence of each outcome [12, 13]. We did not assess the quality of the included primary studies in the SRs nor the quality of reporting of each SR.

We presented summaries of the findings in a format suitable for decision-makers, previously validated during the SUPPORT project [14], focusing on low- and middle-income countries (LMICs) for selected research questions.

Pre-specified subgroups were designated by sampling frame (universal, symptom-based, or risk-based testing), timing of suspicion/diagnosis (pregnancy or postnatal period), trimester of suspicion/diagnosis (first, second or third), country income-level (high or low- and middle-income country), and maternal risk status (low or high).

## Results

The cumulative search retrieval was 608 records, 126 potentially eligible reviews were assessed by full-text and 66 were included for clinical presentation in pregnant women (n=39), maternal outcomes (n=44), clinical presentation in neonates (n=28), neonatal outcomes (n=41) and vertical transmission (n=46) **(Fig 1)**.

**Fig 1.**
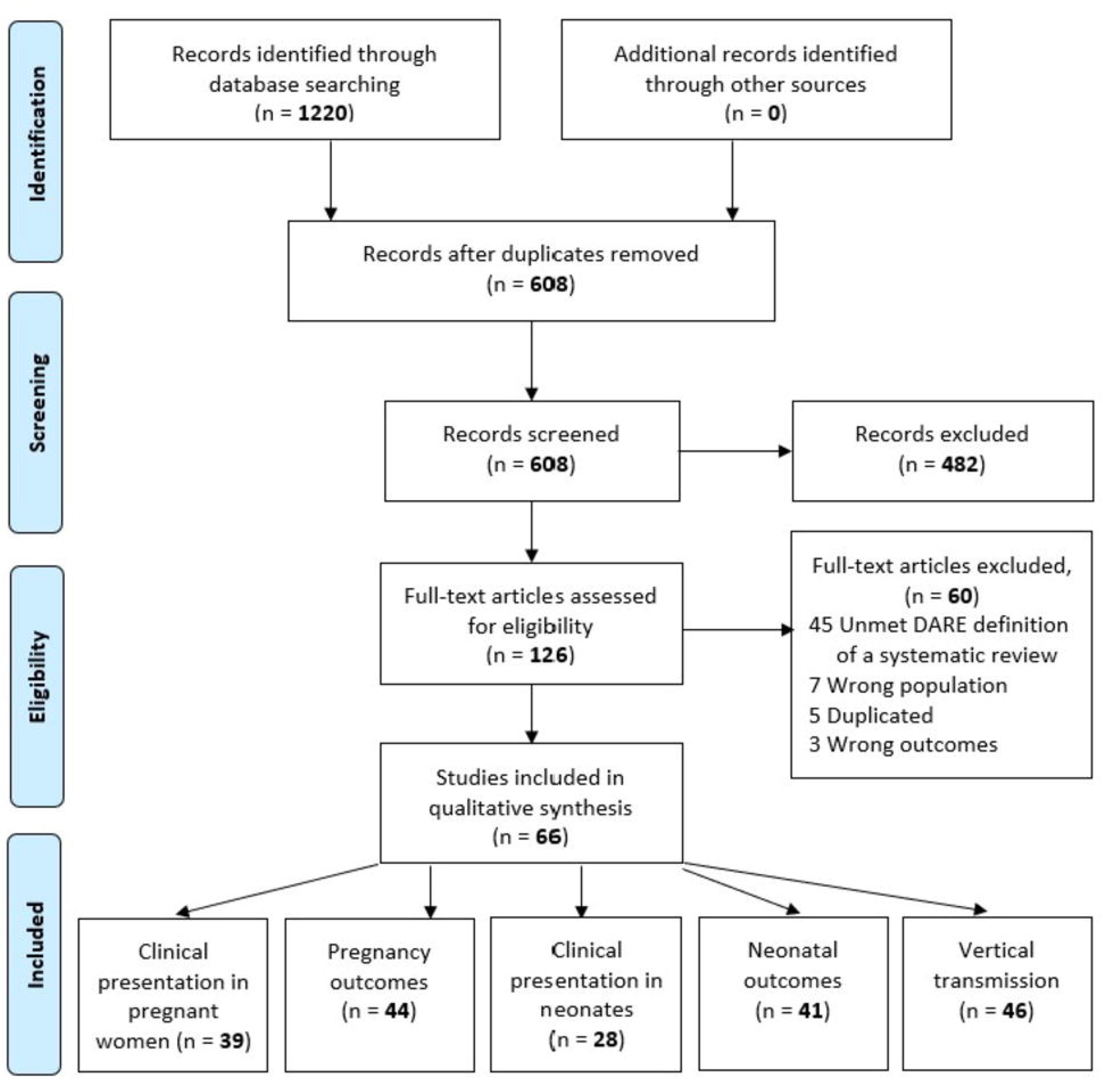
Study flow diagram.

**Table 1** presents the included studies and the list of excluded studies and **S4 Table** the list of excluded studies with their exclusion reasons. All the included SRs were conducted during 2020 all over the world. Although all of them reported a qualitative summary, 18 also included a quantitative summary [2, 15–31].

**Table 1.**
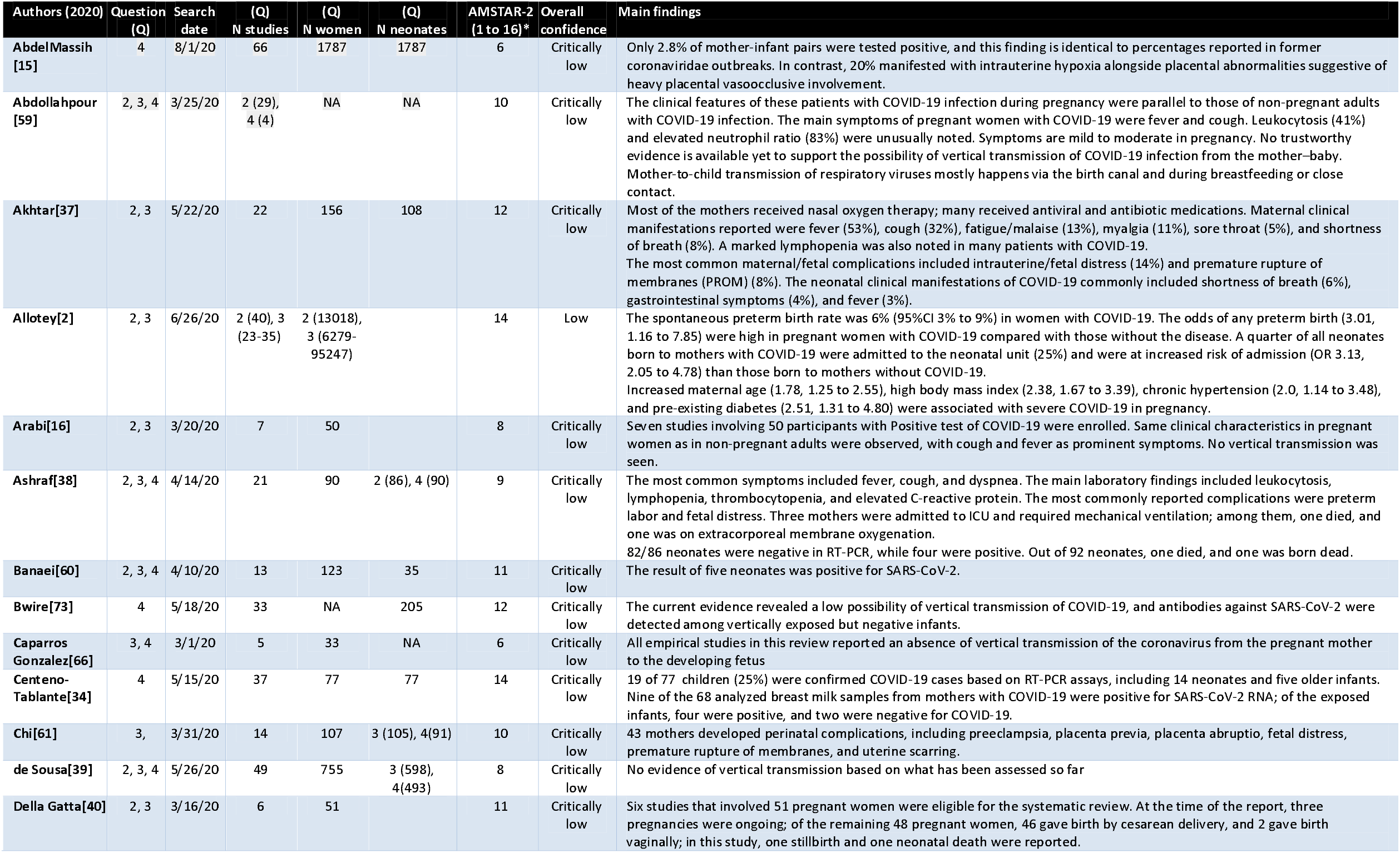
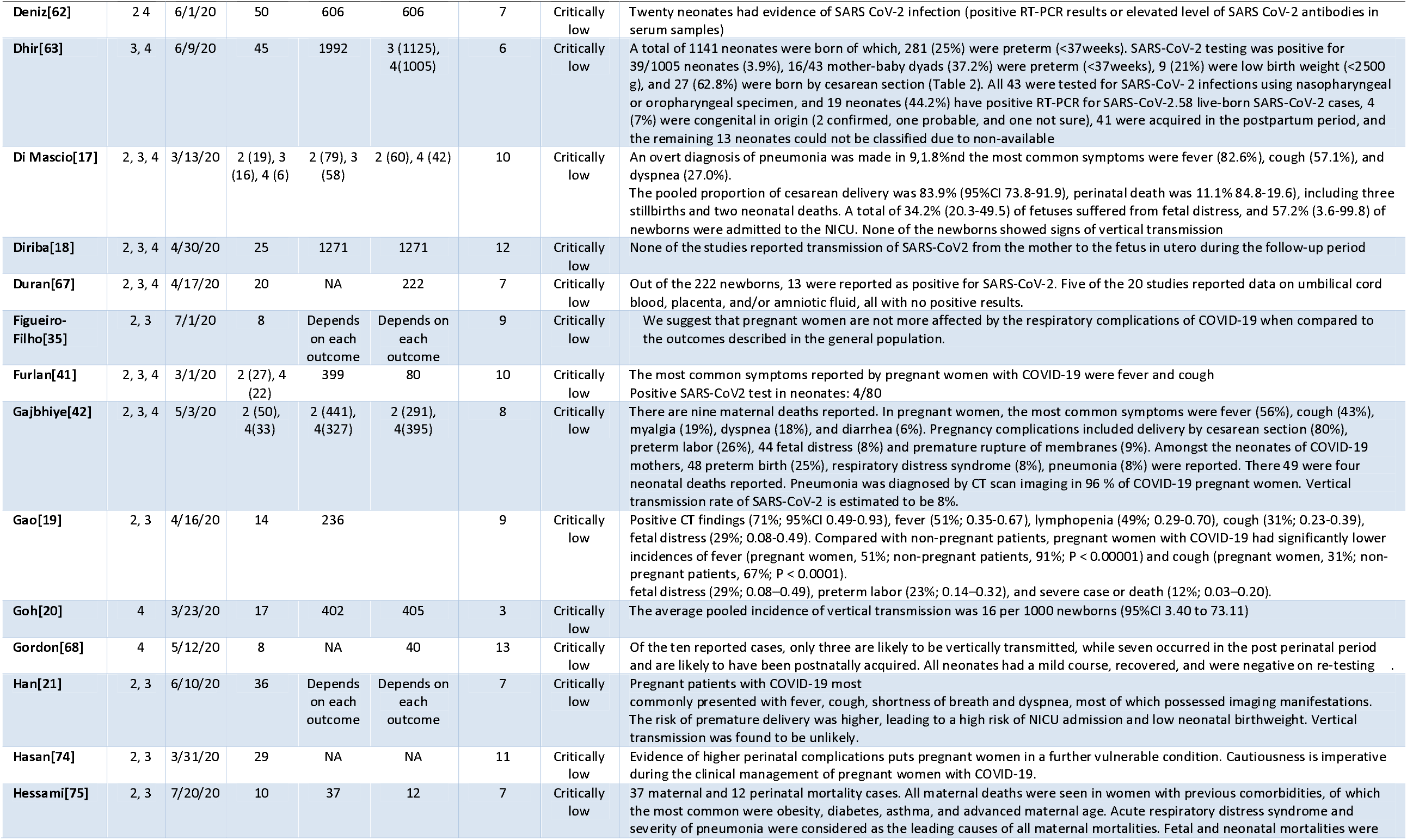
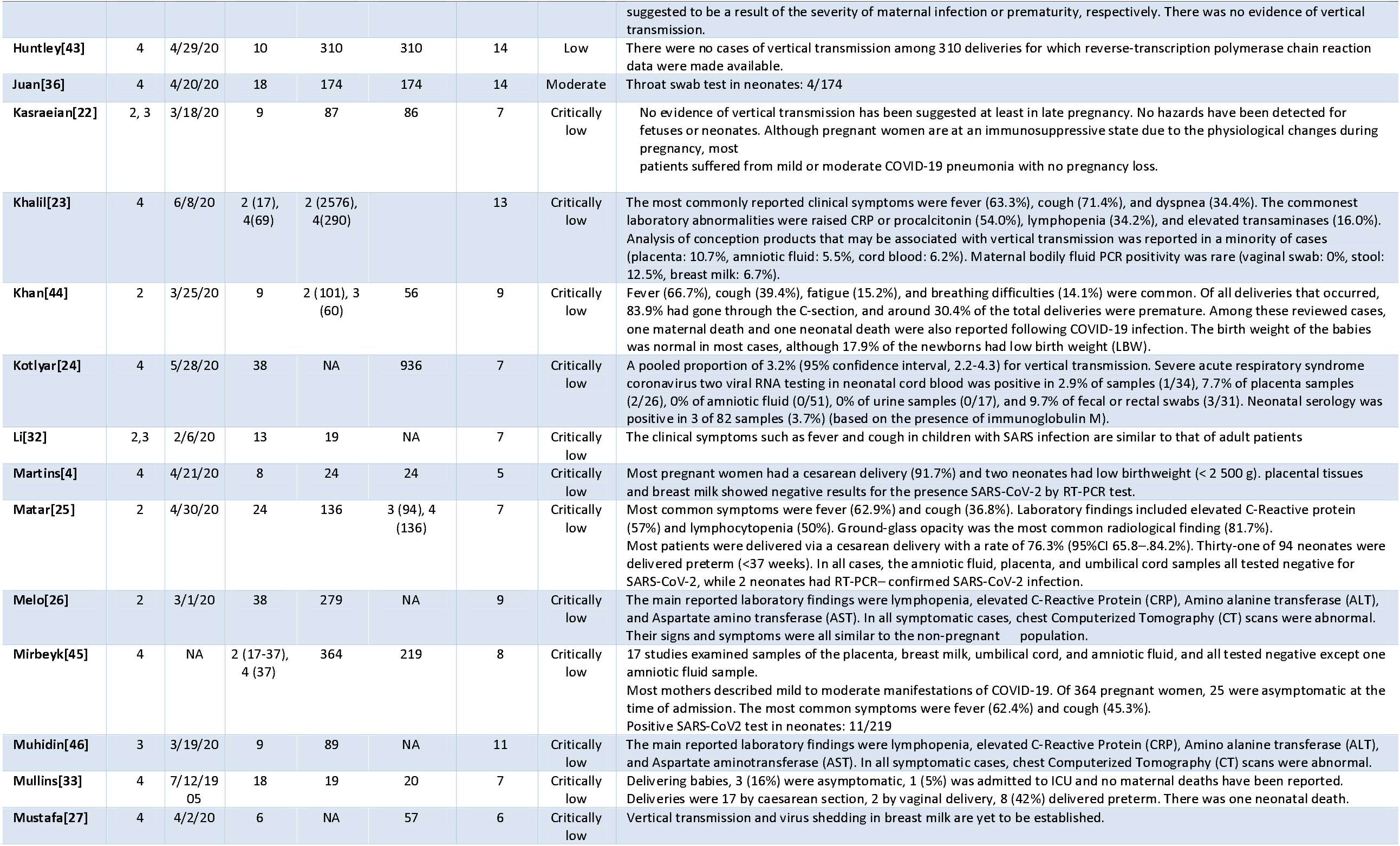
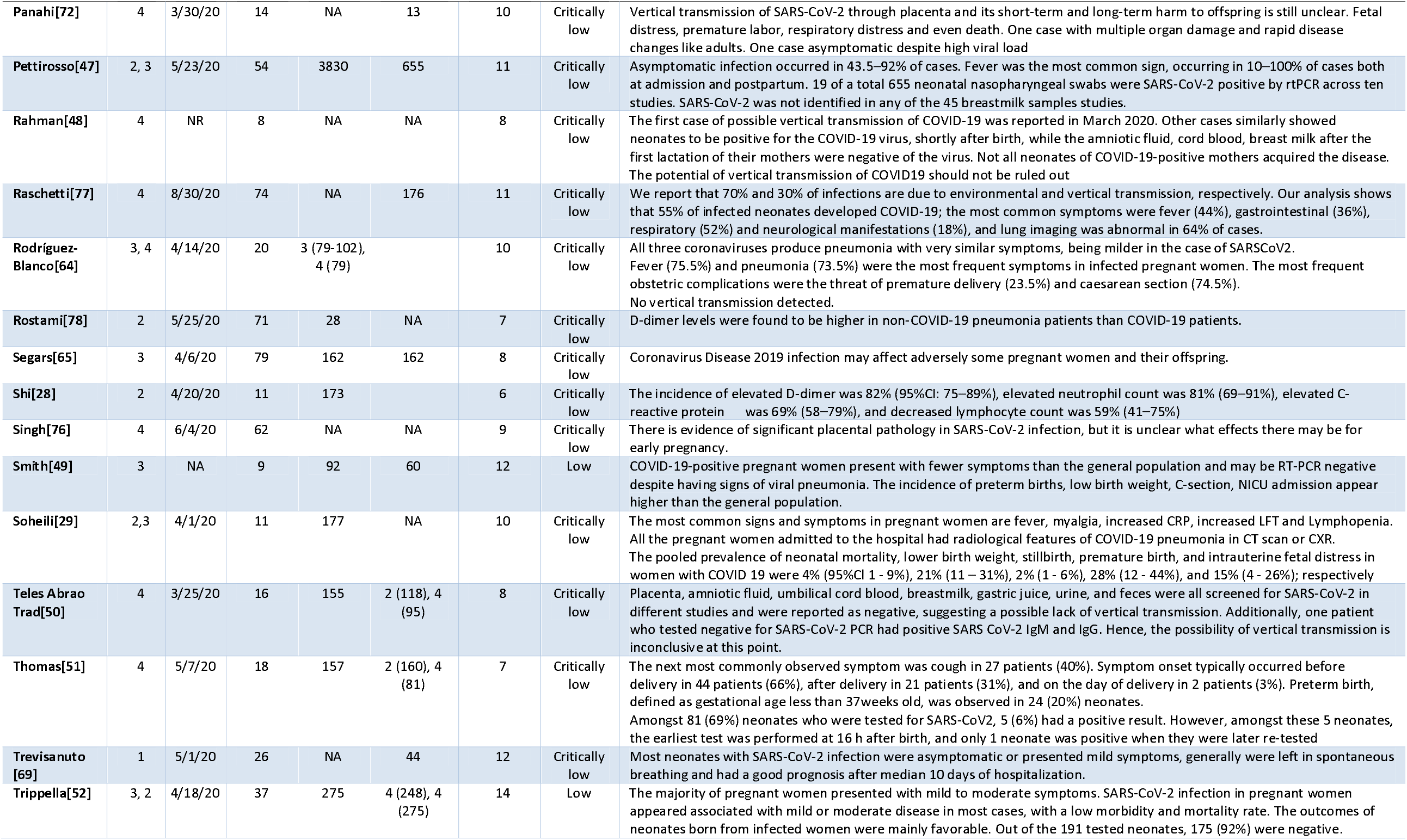
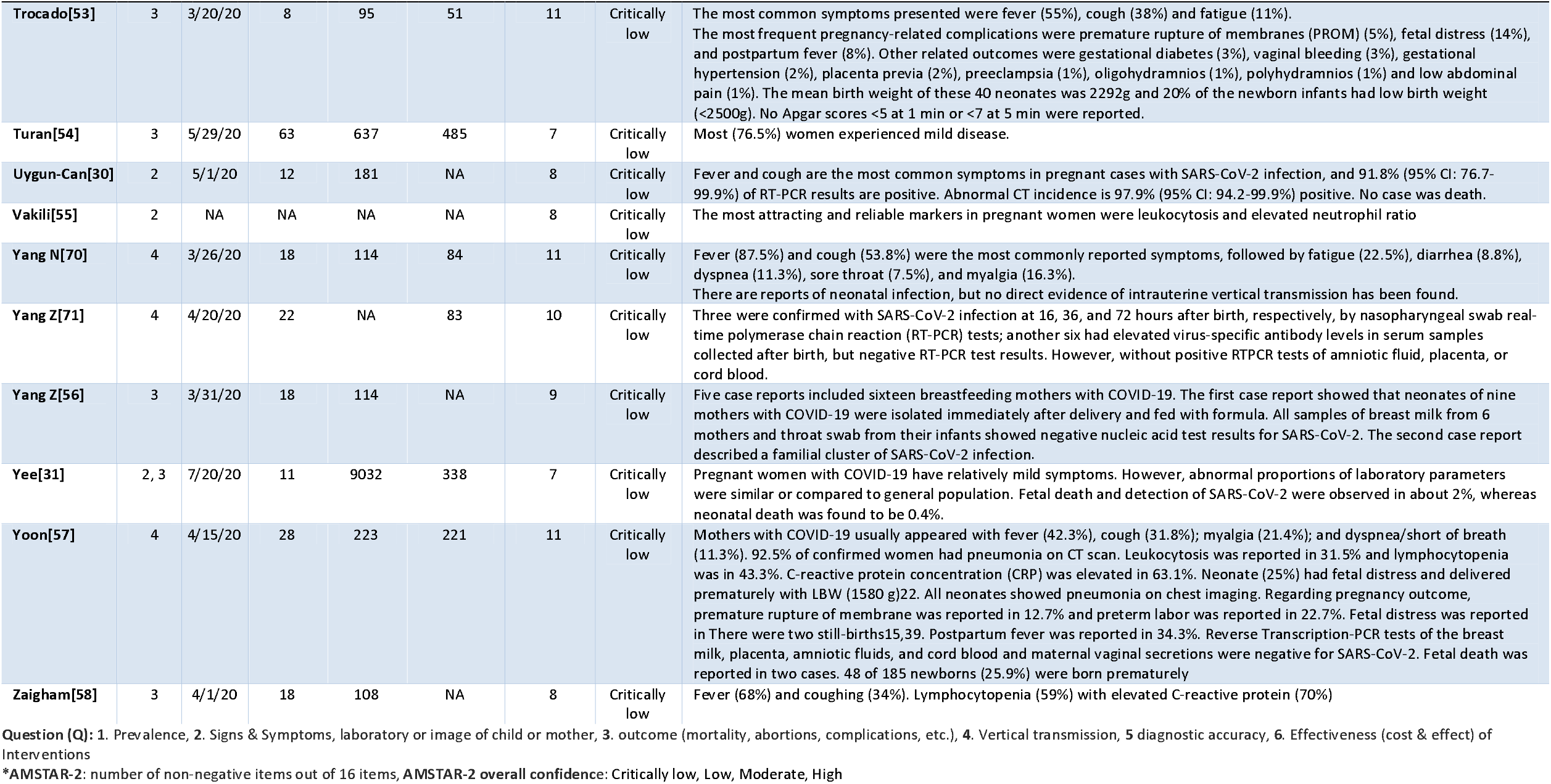
Main characteristics of included systematic reviews.

Among the primary studies included in these reviews, the main study designs were case reports, case series, and other observational studies with or without a comparison group.

The number of included SARS-CoV-2 positive pregnant women was highly heterogeneous across reviews. While the Li review[32] and the Mullins review[33] included only 19 of them, the Allotey review [2] included more than ten thousand.

**Fig 2** shows the percentual degree of overlap of included systematic reviews’ primary studies, which in general was low (In **S5. Primary study list by systematic review** we listed all primary studies with DOI included by each review and in **S6. Primary study overlap matrix across included systematic reviews** the degree of overlap in absolute numbers).

**Fig 2.**
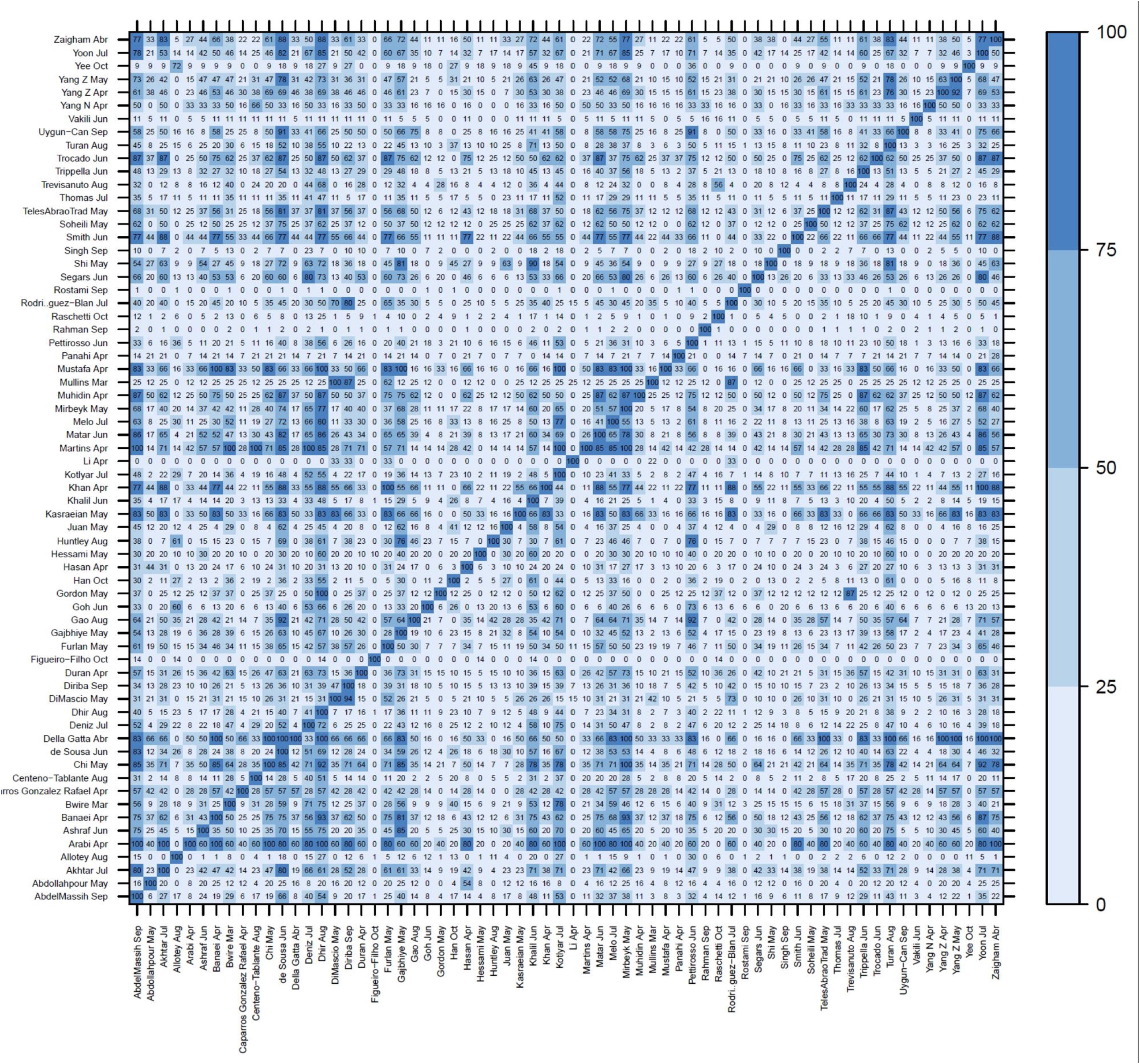
Primary study overlap matrix (percentage) across included systematic reviews*. *The figure displays the 66 included SRs in both axis and the percentage of its primary studies overlap between two SRs. Only primary articles with doi were included for this figure.

Concerning the overall quality, based on AMSTAR-2, most SRs were classified as “critically low” (n=61), four as “low,” and only one as “moderate”. For the prioritized systematic reviews, we also used the ROBIS tool (one was classified as “low risk of bias” [2], one as “unclear risk of bias” [34] and the other two as “high risk of bias” [35, 36]) **(S7. Quality assessment of systematic reviews)**. The mean ± standard deviation of non-negative classifications was 9.21 ± 2.51. The most common weaknesses (>50% of SRs with unmet domain) were: not reporting the funding for the studies included in the review (n=65), not providing a list of excluded studies and justifying the exclusions (n=61), not accounting for risk of bias (RoB) in individual studies when interpreting or discussing the results of the review (n=50), not providing a satisfactory discussion of any heterogeneity observed (n=50), not providing a protocol (n=47) and not using satisfactory techniques for assessing the RoB in individual studies included in the review (n=39).

Below the main findings are described for the five available bodies of evidence identified in our overview: clinical presentation in pregnant women, pregnancy outcomes, clinical presentation in neonates, neonatal outcomes, and vertical transmission.

There was a high level of heterogeneity of the reported values for each outcome across the included SRs. **Table 2** shows these value ranges (The **S8 Systematic review level numerical data by research question** shows the total number of newborns and pregnant women included and the numerical data of each outcome at the review level).

**Table 2.**
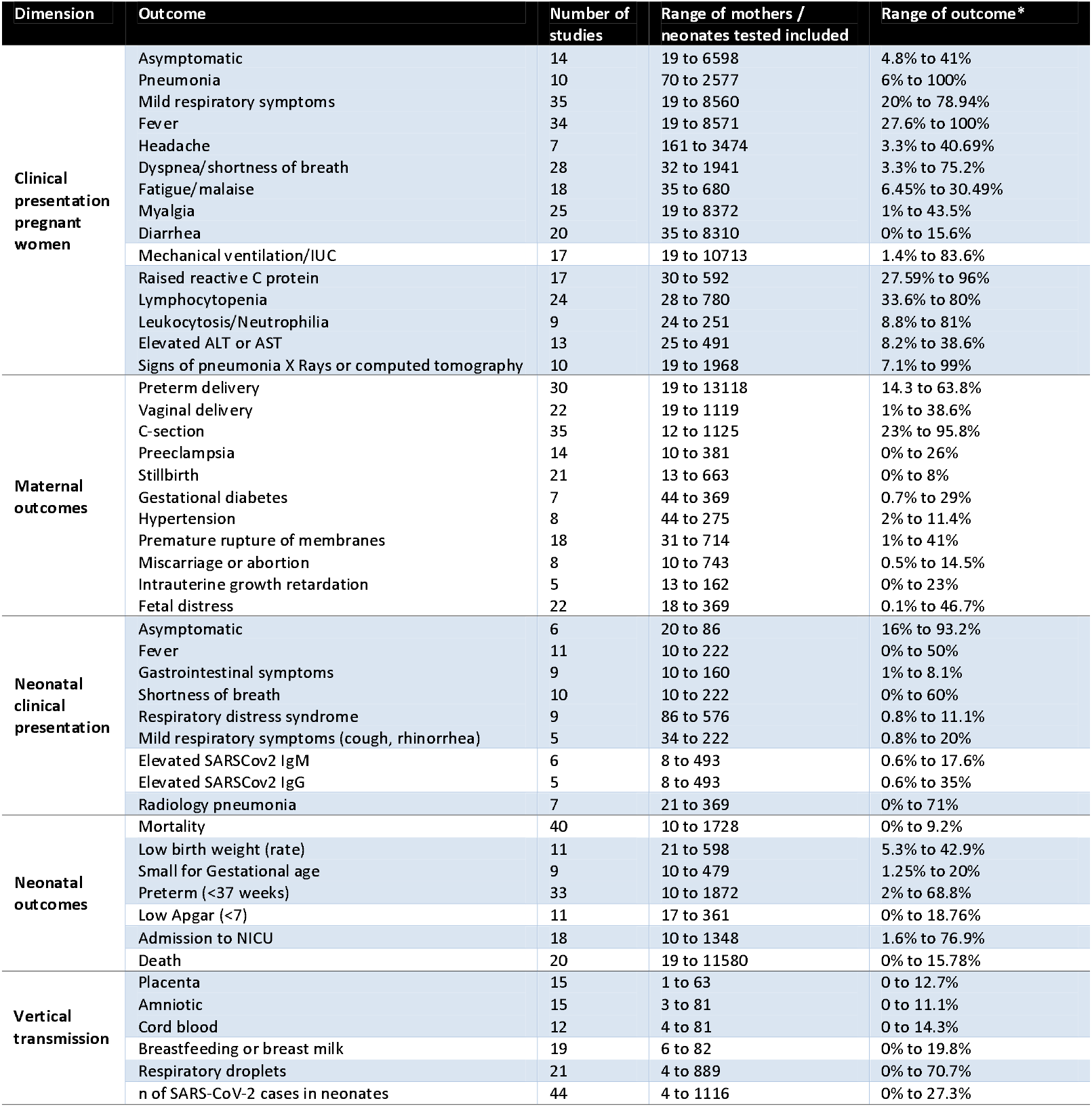
Ranges of outcomes reported in the included systematic reviews.

The main findings are described briefly below.

### Clinical presentation in pregnant women

Thirty-nine reviews[2, 15, 16, 18, 19, 21–23, 25, 28–33, 35–58] showed information regarding clinical manifestations during pregnancy. Fever and mild respiratory symptoms were the most frequently reported symptoms and raised C-reactive protein level, lymphopenia, raised white cell count and raised procalcitonin level were the most frequent laboratory findings. Signs of pneumonia on X-rays or computed tomography (CT) were also frequently reported **(Table 2, S8 Table 1, 2)**.

The Allotey review[2] was chosen as the best review for reporting clinical presentation in pregnant women diagnosed with COVID-19 according to predefined criteria described in the methods section (**Table 3** and **S9 Policy brief)**. It reported that fever (40%), cough (39%, involving 28 studies and 8317 pregnant women), and dyspnea (19%) were the most common symptoms. It also reported that raised C reactive protein levels (49%) and lymphopenia (35%) were the most common laboratory findings. Regarding findings on X-rays or CT, ground glass appearance had a prevalence of 69%, and any other abnormalities on CT had a prevalence of 65%. Finally, when compared to non-pregnant women of reproductive age with COVID-19, pregnant women with the disease were less likely to manifest fever (OR 0.43, 95%CI 0.22-0.85) and myalgia (OR 0.48, 95%CI 0.45-0.51).

**Table 3.**
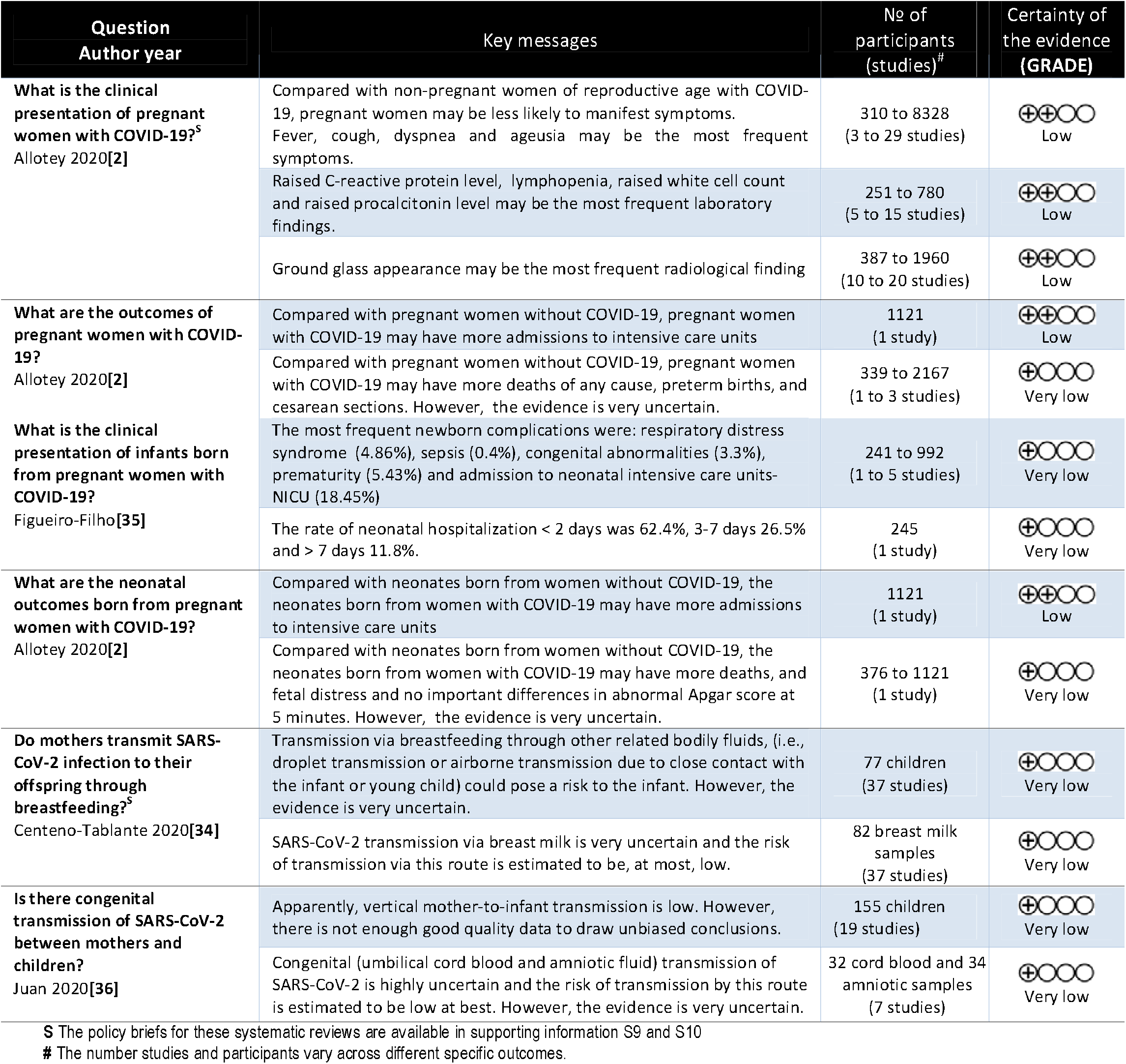
Key messages of prioritized systematic reviews.

| Question<br>Author year | Key messages | Nº of<br>participants<br>(studies)<sup>#</sup> | Certainty of<br>the evidence<br>(GRADE) |
| --- | --- | --- | --- |
| **What is the clinical presentation of pregnant women with COVID-19?**<sup>S</sup><br>Allotey 2020[2] | Compared with non-pregnant women of reproductive age with COVID-19, pregnant women may be less likely to manifest symptoms. Fever, cough, dyspnea and ageusia may be the most frequent symptoms. | 310 to 8328<br>(3 to 29 studies) | ⊕⊕○○<br>Low |
|  | Raised C-reactive protein level, lymphopenia, raised white cell count and raised procalcitonin level may be the most frequent laboratory findings. | 251 to 780<br>(5 to 15 studies) | ⊕⊕○○<br>Low |
|  | Ground glass appearance may be the most frequent radiological finding | 387 to 1960<br>(10 to 20 studies) | ⊕⊕○○<br>Low |
| **What are the outcomes of pregnant women with COVID-19?**<br>Allotey 2020[2] | Compared with pregnant women without COVID-19, pregnant women with COVID-19 may have more admissions to intensive care units | 1121<br>(1 study) | ⊕⊕○○<br>Low |
|  | Compared with pregnant women without COVID-19, pregnant women with COVID-19 may have more deaths of any cause, preterm births, and cesarean sections. However, the evidence is very uncertain. | 339 to 2167<br>(1 to 3 studies) | ⊕○○○<br>Very low |
| **What is the clinical presentation of infants born from pregnant women with COVID-19?**<br>Figueiro-Filho[35] | The most frequent newborn complications were: respiratory distress syndrome (4.86%), sepsis (0.4%), congenital abnormalities (3.3%), prematurity (5.43%) and admission to neonatal intensive care units- NICU (18.45%) | 241 to 992<br>(1 to 5 studies) | ⊕○○○<br>Very low |
|  | The rate of neonatal hospitalization < 2 days was 62.4%, 3-7 days 26.5% and > 7 days 11.8%. | 245<br>(1 study) | ⊕○○○<br>Very low |
| **What are the neonatal outcomes born from pregnant women with COVID-19?**<br>Allotey 2020[2] | Compared with neonates born from women without COVID-19, the neonates born from women with COVID-19 may have more admissions to intensive care units | 1121<br>(1 study) | ⊕⊕○○<br>Low |
|  | Compared with neonates born from women without COVID-19, the neonates born from women with COVID-19 may have more deaths, and fetal distress and no important differences in abnormal Apgar score at 5 minutes. However, the evidence is very uncertain. | 376 to 1121<br>(1 study) | ⊕○○○<br>Very low |
| **Do mothers transmit SARS-CoV-2 infection to their offspring through breastfeeding?**<sup>S</sup><br>Centeno-Tablante 2020[34] | Transmission via breastfeeding through other related bodily fluids, (i.e., droplet transmission or airborne transmission due to close contact with the infant or young child) could pose a risk to the infant. However, the evidence is very uncertain. | 77 children<br>(37 studies) | ⊕○○○<br>Very low |
|  | SARS-CoV-2 transmission via breast milk is very uncertain and the risk of transmission via this route is estimated to be, at most, low. | 82 breast milk samples<br>(37 studies) | ⊕○○○<br>Very low |
| **Is there congenital transmission of SARS-CoV-2 between mothers and children?**<br>Juan 2020[36] | Apparently, vertical mother-to-infant transmission is low. However, there is not enough good quality data to draw unbiased conclusions. | 155 children<br>(19 studies) | ⊕○○○<br>Very low |
|  | Congenital (umbilical cord blood and amniotic fluid) transmission of SARS-CoV-2 is highly uncertain and the risk of transmission by this route is estimated to be low at best. However, the evidence is very uncertain. | 32 cord blood and 34 amniotic samples<br>(7 studies) | ⊕○○○<br>Very low |

<sup>S</sup> The policy briefs for these systematic reviews are available in supporting information S9 and S10
<sup>#</sup> The number studies and participants vary across different specific outcomes.

### Maternal outcomes

Forty-four reviews[2, 4, 15–19, 21–23, 25, 26, 29, 31–33, 35–38, 40–47, 49–54, 56–65] reported at least one pregnancy outcome in pregnant women. The main findings are presented in **Table 3, S8 Table 3**. The most frequently reported outcomes were C-section and preterm delivery. The Allotey review[2] was chosen as the best review for reporting maternal outcomes according to predefined criteria **(Table 3)**. It was reported that the prevalence of all-cause mortality was 0.63%, severe COVID-19 was 13%, admission to an intensive care unit (ICU) was 4%, and required invasive ventilation was 3%.

When compared with non-pregnant women of reproductive age with COVID-19, the reported odds of admission to the ICU was 1.62 (95%CI 1.33-1.96); and the reported odds of required invasive ventilation was 1.88 (95%CI 1.36-2.60).

Maternal risk factors associated with severe COVID-19 were age (OR 1.78, 95%CI 1.25-2.55), high body mass index (OR 2.38, 95%CI 1.67-3.39), hypertension (OR 2.0, 95%CI 1.14-3.48), and pre-existing diabetes (OR 2.51, 95%CI 1.31-4.80).

### Neonatal clinical presentation

Twenty-eight reviews[2, 15, 31, 33, 35–39, 42, 45, 46, 49–54, 57, 60, 61, 63, 66–71] reported information on the clinical presentation in neonates born from pregnant women diagnosed with SARS-CoV-2 **(Table 2, S8 Table 4)**.

Respiratory distress syndrome, shortness of breath, and fetal distress were the most frequently reported moderate to severe presentations.

Laboratory data (confirmed cases through Polymerase chain reaction [PCR], elevated SARS-CoV-2 IgM, and IgG antibodies) and imaging data (radiographic pneumonia) were also reported.

The Figueiro-Filho review [35] was chosen as the best review for reporting neonatal clinical presentation according to predefined criteria **(Table 3)**. This review involved almost 11,000 cases of COVID-19 and pregnancy in 15 different countries. The most frequent newborn complications were admission to neonatal ICUs (NICUs) (18.5%), prematurity complications (5.43%), respiratory distress syndrome (4.9%), and congenital abnormalities (3.3%). The rate of neonatal hospitalization < 2 days was 62.4% and > 7 days 11.8%.

### Neonatal outcomes

Forty-one studies[2, 16–19, 21–23, 25, 29, 31, 33, 35–38, 40–43, 45, 46, 49–54, 57, 58, 60–67, 69, 70, 72] presented neonatal outcomes of pregnant women diagnosed with SARS-CoV-2 **(Table 2, S8 Table 5)**. The most frequently reported neonatal outcomes were low birth weight and preterm delivery.

The Allotey review[2] was chosen as the best review for reporting neonatal outcomes according to predefined criteria **(Table 3)**. The reported neonatal mortality prevalence was 0.34%, and neonatal admission to NICUs was 25% (95%CI 14%-37%). The neonates born to mothers diagnosed with COVID-19 presented an odds ratio of admission to critical care unit equal to 3.13 (95%CI OR 2.05-4.78) compared with those born to mothers without the disease. Finally, the rate of overall preterm birth reported among pregnant women diagnosed with COVID-19 was 17% (95%CI 13%-21%). No significant findings were observed for other neonatal outcomes.

Vertical transmission

Forty-six reviews[4, 15, 17, 18, 20, 21, 23–27, 31, 33–36, 38, 39, 41, 42, 45, 47–52, 54, 56–64, 66–68, 70, 71, 73–76] reported mother-to-child SARS-CoV-2 transmission **(Table 2, S8 Table 6)**. Most studies only reported the proportion of infants’ positive cases without evaluating breast milk or congenital/perinatal transmission. In the reviews reporting breast milk or congenital/perinatal transmission the sample analyzed was generally small for these outcomes.

The Centeno-Tablante review[34] was chosen as the most appropriate review to answer this question according to predefined criteria **(Table 3** and **S10 Policy brief)**. This review included 37 papers with a total of 889 infants. Of the 72 infected mothers whose breast milk samples were laboratory-confirmed to contain the COVID-19 antigen, 14 infants were found to be infected with COVID-19. Eight of the twenty-three infants that were breastfed were infected, two of the eighteen infants that received a breast milk substitute were infected, two of four infants that received mixed feeding were infected, and two of the twenty-three infants that did not report on feeding practice were infected. Regarding congenital/perinatal transmission cases, the Juan review[36] was deemed to be the most appropriate review **(Table 3)**. This review included 24 studies, case series, and case reports, including a total of 155 neonates. Ninety neonates were tested for COVID-19, of which three were positive. The review also evaluated the presence of SARS-CoV-2 in amniotic fluid (1/32), umbilical cord blood (0/34), and placenta (1/3). While there was no vertical mother-to-child transmission, additional good-quality studies are needed to determine whether vertical transmission is possible.

The key messages from the prioritized systematic reviews (by the most current search date, the more significant number of included studies, and greater adequacy to address the outcomes) are presented in **Table 3**.

## Discussion

This systematic review of SRs integrated the most consolidated evidence synthesis regarding the effects of COVID-19 on maternal and neonatal health.

Most SRs (92.4%) were classified as “critically low” in overall confidence, using the AMSTAR-2 tool, likely due to the urgent demand of information for this hot topic. For the prioritized systematic reviews, we also used the ROBIS tool (one was classified as “low risk of bias” [2], one as “unclear risk of bias” [34] and the other two as “high risk of bias” [35, 36]) and the GRADE approach for each of their outcomes. The certainty of evidence was rated as “low” to “very low” due to study design, risk of bias, inconsistency and/or imprecision.

The COVID-19 related symptoms manifest from one third to two thirds less often in pregnant women than in non-pregnant women of reproductive age [2]. While testing for SARS-CoV-2 in non-pregnant women is usually based on symptoms or contact history, testing in pregnant women is usually done for reasons that might not be related to COVID-19. Pregnant women with COVID-19 seem to be at increased risk of requiring admission to an ICU or invasive ventilation. Pregnant women with COVID-19 are also at an increased risk of receiving cesarean sections, delivering preterm and their babies being admitted to a NICU. Increased maternal age, high body mass index, and pre-existing comorbidities might be associated with severe disease [2]. Stillbirth and neonatal death rates are low in women with suspected or confirmed COVID-19. All comparative findings are based on a small number of studies, despite the large sample sizes. Substantial heterogeneity was observed in the estimates for rates of clinical manifestations and outcomes, which varied by sampling frames, participant selection, and risk status of the participants.

Most infected neonates were asymptomatic. If clinical manifestation was present, the most frequent symptoms were fever (0-50%) or mild respiratory symptoms [35]. Low birth weight and preterm birth were the most frequently reported neonatal outcomes. The neonates born to mothers diagnosed with COVID-19 presented an odds three times higher of admission to a NICU compared with those born to mothers without the disease [2]. The risk of congenital transmission [36] or transmission via breast milk [34] is estimated to be low to very low, but there is higher risk of transmission due to close contact by droplet or airborne transmission.

As expected, high rates of asymptomatic presentation in pregnant women with COVID-19 were found [2]. This is likely because of the strategy of universal screening for COVID-19 in pregnancy and the low thresholds for testing in non-pregnancy. Despite the possibility of the above strategies detecting pregnant women with mild disease, admissions to the ICU and the need for invasive ventilation was observed when compared with non-pregnant women of reproductive age with COVID-19 [79]. The pooled estimates for severe COVID-19 and admission to an ICU were, however, still relatively high in the non-comparative data, indicative of a potentially high risk in pregnancy. This is also supported by the recent analysis in a Swedish study suggesting an increased risk of admission to an ICU and of invasive ventilation in pregnant women than non-pregnant women [80].

Similar to the general population, pre-existing comorbidities seemed to be risk factors for severity of COVID-19 in pregnancy [81]. Complications related to COVID-19 did not seem to be increased in women presenting in the third trimester or in multiparous women—but the existing sample sizes are not large (less than 300 women). Both chronic hypertension and pre-existing diabetes were associated with maternal death in pregnant women with COVID-19, both of which are known risk factors in the general population. It is not known if COVID-19 was the direct cause of death for these women, and the numbers of studies were small. A slight increase in rates of preterm birth in pregnant women with COVID-19 was observed when compared to those without the disease. These preterm births could be medically indicative, as the overall rates of spontaneous preterm births in pregnant women with COVID-19 was broadly similar to those observed in the pre-pandemic period. More than 60% of pregnant women underwent cesarean section in the non-comparative studies. This is three times the global rate of cesarean sections worldwide [82], and deserves future research.

The precision of the estimates is expected to improve as more data is published. The overall rates of stillbirths and neonatal deaths are not observably higher than the background rates. The indicators for admissions to the NICU, observed in about a quarter of neonates delivered to mothers with COVID-19, have not been reported. Local policies on observation and quarantine of infants with exposure to SARS-CoV-2 may have influenced these rates.

Sixty-seven percent of newborns delivered by mothers with COVID-19 antibodies had SARS-CoV-2 IgG, but not IgM antibodies [83]. This finding against vertical transmission is consistent with our own findings of low to very low risk of this mechanism of congenital transmission.

To our knowledge, there is only one overview published that reports maternal and perinatal outcomes related to COVID-19 and pregnancy [84], including 52 SRs. This overview searched studies until September 2020 and did not include 14 SRs that were found in our overview, probably explaining the lower level of overlap observed in our study. The authors did not assess the quality of reporting of each SR, but assessed the risk of bias of each included SR using the ROBIS tool [11]. The high risk of bias identified in this overview is consistent with the “critically low” confidence presented in our study by applying the AMSTAR-2 tool [10].

Alongside the spread of the pandemic, a shift has occurred in the types of studies published, with initial studies involving pregnant women from epidemic regions in China, followed by reports of large regional and national datasets from the US, UK, Netherlands, Spain, and, more recently, Latin American countries. The study design has also changed from small case series and case reports to extensive observational data, with recent studies also providing comparative data. The prevalence of COVID-19 varied widely between studies, particularly when sampling was done based on symptoms or history of contact, highlighting the variations in criteria for testing.

Moreover, the findings only apply to women attending the hospital for any reason. The true prevalence of COVID-19 in pregnancy is likely to be lower when all pregnant women are included.

### Strengths and limitations

This overview has several strengths. First, we followed sound methodology to conduct the present overview of systematic reviews. Second, we included systematic reviews without language restrictions. Third, we adhered to rigorous quality appraisal for the conduction of systematic reviews (AMSTAR-2 tool), which was independently assessed by two reviewers and discrepancies solved by consensus. We summarized and critically appraised an important amount of evidence that is relevant to health decision-making (See examples of policy briefs in supporting information S9 and S10) and highlighted evidence gaps that could guide future research. Finally, we conducted a sensitive and comprehensive search strategy to reduce the risk of missing relevant studies. We synthesized the results of all relevant SRs, highlighting their overlap through a matrix of primary studies by SR, and we also selected the best SR that answered a specific question according to pre-defined criteria of relevance, comprehensiveness, data update, and quality. We presented these SRs through a summary of the findings tables with the certainty of evidence according to the GRADE approach. Our review integrated the evidence generated by different independent groups, which could improve the robustness of our findings.

Our study is not exempt from limitations. A main limitation is that the last search was run in October 2020. This is due to the time needed to perform a through SR. The general confidence of the included SRs was “critically low”, and the certainty of evidence was “low” to “very low”. Additionally, there is a scarcity of data comparing pregnant women with non-pregnant women or comparing pregnant women with and without COVID-19 [2]. We did not assess the risk of bias of the primary studies, nor did we undertake a pooled analysis by the outcome, as was originally stated in our protocol. Nevertheless, there was great heterogeneity of methods, study designs, and estimations that would preclude a meta-analysis.

### Implications for clinical practice and research

Based on existing data, healthcare professionals should be aware that pregnant women with COVID-19 might manifest fewer symptoms than the general population, though the overall pattern is similar. Emerging comparative data indicate the potential for an increase in the rates of admission to ICUs and invasive ventilation in pregnant women with COVID-19 compared with infected non-pregnant women. Mothers with pre-existing comorbidities will need to be considered as a high-risk group for COVID-19, along with those who are obese and of advanced maternal age. Clinicians will need to balance the need for regular multidisciplinary antenatal care to manage women with pre-existing comorbidities against unnecessary exposure to the virus through virtual clinic appointments when possible. Pregnant women with COVID-19 before term gestation might need to be managed in a unit with facilities to care for preterm neonates since the neonates born to mothers diagnosed with COVID-19 are at three times the risk of admission to NICUs compared with those born to mothers without the disease.

Further data are needed to assess robustly if pregnancy-related maternal and neonatal complications are increased in women with COVID-19 compared to those without the disease. Similarly, the association between other risk factors, such as ethnicity and pregnancy-specific risk factors such as preeclampsia and gestational diabetes on both COVID-19 related and pregnancy-related outcomes needs evaluation. Preeclampsia was reported to be associated with severe COVID-19 in small studies, but requires further assessment as the clinical presentation of severe preeclampsia could mimic worsening COVID-19 [85]. Robust collection of maternal data by trimester of exposure, including the periconception period, is required to determine the effects of COVID-19 on early pregnancy outcomes, fetal growth, and risk of stillbirth.

Systematic reviews are considered to be the highest quality evidence to inform guidelines, and so poor-quality systematic reviews will still have a direct impact on clinical care. Despite the urgent need for evidence on the impact of COVID-19 in pregnant women, systematic reviews and meta-analyses still need to adhere to the reporting guidelines on search criteria, quality assessment, and analysis. This is particularly important as large numbers of non-peer-reviewed scientific papers and reports are currently available in the public domain in multiple versions. Primary studies will need to explicitly state if duplicate data have been included to avoid double counting of participants in evidence synthesis. Individual participant data meta-analysis and network meta-analysis of the emerging cohorts are critical to assess both differential presentation and outcomes by underlying risk factors and to determine the differential effects of interventions to reduce the rates of complications.

## Data Availability

All relevant data are within the manuscript and its Supporting Information files.

## Author contributions

AC, AB, MB, AM and PB contributed to conception and design of this manuscript and prepared the first draft. All authors interpreted the data, revised critically the first draft and signed off on the final version.

## Acknowledgments

We thank Erin Goucher, for her help with English edition of the manuscript.

## Supporting information

**S1**. Preferred Reporting Items for Overviews of systematic reviews

**S2**. Outcomes of interest

**S3**. Search strategy

**S4**. Excluded studies & exclusion reasons

**S5**. Primary study list by systematic review

**S6**. Primary study overlap matrix, in absolute numbers, across included systematic reviews

**S7**. Quality assessment of systematic reviews

**S8**. Extracted data at systematic review level by the research question

**S9**. Policy brief of clinical presentation of pregnant women with COVID-19

**S10**. Policy brief of transmission of SARS-CoV-2 through breastfeeding

