## Supplementary material for "COVID-19 and pregnancy: An umbrella review of clinical presentation, vertical transmission, and maternal and perinatal outcomes": S1 to S10: S1. Preferred Reporting Items for Overviews of systematic reviews.docx

### S1. Preferred Reporting Items for Overviews of systematic reviews including harms checklist

| **Section/Topic** | **(Sub-) item #** | **Checklist item** | | | | | **Page #** |
| --- | --- | --- | --- | --- | --- | --- | --- |
| **TITLE** | | | | | | |  |
| 1. Title | 1a | Specify the study design with terms such as “overview of (systematic) reviews,” “umbrella review,” “(systematic) review of systematic reviews,” or “(systematic) meta-review” in the title of the OoSRs. | | | | | 1 |
|  | 1b | Mention “safety” or harms related terms, or the adverse event(s) of interest in the title of the OoSRs. | | | | | 2, 5 |
| **ABSTRACT** | | | | | | |  |
| 2. Structured-like summary | 2a | Provide a structured-like abstract, as applicable: background, objective, data sources, selection criteria, data extraction, review appraisal, data synthesis methods, results, limitations, conclusions. | | | | | 1 |
|  | 2b | Report the main findings of analysis of harms undertaken in the OoSRs or/and in the included SRs. | | | | | 1 |
| **INTRODUCTION** | | | | | | |  |
| 3. Rationale | 3a | Specify the rationale and the scope (wide or narrow agendas) for the overview in the context of an existing body of knowledge on the topic. | | | | | 2 |
|  | 3b | Provide a balanced presentation of potential benefits and harms of the intervention(s). | | | | | 2 |
|  | 3c**^a^** | Define which events are considered harms according to previous literature and provide a clear rationale for the specific harms included in the OoSRs. | | | | | 2 |
| 4. Objectives  (PICOS) | 4 | Provide an explicit statement of research question(s) that specifies PICOS: | | | | | 3 |
|  |  | Participants 🗷 | Interventions 🗷 | Comparators 🗷 | Outcomes 🗷 | Study design🗷 |  |
| **METHODS** | | | | | | |  |
| 5. Protocol and registration | 5a | Indicate clearly if a protocol exists or not. | | | | | 3 |
|  | 5b | If registered, provide the name of the registry (such as a valid Web address, PROSPERO). | | | | | 3 |
| 6. Eligibility criteria  & outcomes of interest | 6a | Specify inclusion and exclusion criteria for study design, participants, interventions and comparators in detail. | | | | | 3 |
|  | 6b | List (and define whenever it is necessary) the outcomes for which data were recorded, ideally include prioritization of main and additional outcomes. | | | | | 3, S2 |
|  | 6c | Include adverse events as (primary or secondary) outcome of interest. Define them and grade their severity (such as mild, moderate, severe, fatal; severity could also be described in the appendix), if appropriate. | | | | | 3, S2 |
|  | 6d**^b^** | Specify report characteristics (such as language restrictions, publication status, and years considered) used as criteria for eligibility for the OoSRs (see also item 7). | | | | | 3 |
| 7. Information sources | 7a | Search at least two electronic bases. | | | | | 3 |
|  | 7b | Search supplementary sources (e.g. hand-searching, reference lists, related reviews and guidelines, protocol registries, conference abstracts, and other gray literature). | | | | | 3 |
|  | 7c | Report the date last searched and/or dates of coverage for each database. | | | | | 3 |
| 8. Search strategy**^c^** | 8a | Specify full electronic search strategy (algorithm) for at least one database including any limits used (e.g. language and date restrictions-see also subitems 6d and 7c) such that it could be repeated. | | | | | S3 |
|  | 8b | Present any additional search process (e.g. algorithm or filter for adverse events, searches in pertinent websites) specifically to identify adverse events that have been investigated. | | | | | - |
| 9. Data management &  selection process | 9a**^d^** | Describe the software that was used to manage records and data throughout the OoSRs. | | | | | 3 |
|  | 9b | Define what is a SR and provide the process for selecting SRs and its relevant details (screening the title and abstract or full text by at least two reviewers, selection by multiple independent investigators and resolving disagreements by consensus). | | | | | 3 |
|  | 9c | Report any attempt to handle overlapping (include one review among multiple potential candidates by choosing for example the most updated SR, the most methodologically rigorous SR or the SR with larger number of primary studies). | | | | | 3,4 |
| 10. Additional search for  primary studies | 10 | Report additional search to identify eligible primary studies (e.g. searching in more databases or update the search) and its relevant details. | | | | | 3 |
| 11. Data collection process | 11a | Describe the method of data extraction from included SRs (e.g. data collection form, extraction in duplicate and independently, resolving disagreements by consensus). | | | | | 3 |
|  | 11b | Report any processes for obtaining, confirming or updating data from investigators (e.g. contact with authors of included reviews, obtain data from primary studies of included reviews). | | | | | 3 |
| 12. Data items | 12 | List (and define whenever is necessary) the specific variables for which data were recorded (e.g. PICOS items, number of included studies and participants, dose, length of follow up, results, funding sources) and any data assumptions and simplifications made. | | | | | 3, 4 |
| 13. Assessment of methodological quality & quality of evidence | 13a | State the evaluation of reporting or/and methodological quality (eg. using PRISMA or PRISMA-harms, AMSTAR or R-AMSTAR) of the included reviews. | | | | | 4 |
|  | 13b**^e^** | State the evaluation of quality for individual studies that were included in the SRs (inform whether tools such as Jadad or RoB of Cochrane were used by the included reviews) and for the additional primary studies. | | | | | NA |
|  | 13c | State the evaluation of quality of evidence (e.g. using GRADE approach). | | | | | Table 3 |
|  | 13d | Describe the methods (e.g. piloted forms, independently, in duplicate) used for the quality assessment. | | | | | 4 |
| 14. Meta-bias(es) | 14 | Specify any planned assessment of meta-bias(es) (such as publication bias or selective reporting across studies, ROBIS tool). | | | | | NA |
| 15. Data synthesis | 15a | Specify clearly the method (narrative, meta-analysis or network meta-analysis) of handling or synthesizing data and their details (e.g. state the principal summary measures that were extracted or calculated, how heterogeneity was assessed, what statistical approaches were used if a quantitative synthesis has been conducted). | | | | | 4 |
|  | 15b | Describe the software that was used to analyze the data if a quantitative synthesis has been conducted. | | | | | NA |
|  | 15c | Report if zero events are included in the studies and how they were handled in statistical analyses, if relevant. | | | | | NA |
|  | 15d | Describe methods of any pre-specified additional analyses (such as sensitivity or subgroup analyses, meta-regression). | | | | | NA |
| **RESULTS** | | | | | | |  |
| 16. Review & primary study selection | 16a | Provide the details of review selection (e.g. numbers of reviews screened, retrieved, and included and excluded in the overview) and the number of the additional eligible primary studies that were included, ideally with a flow diagram of the overview process. | | | | | 4, Fig 1 |
|  | 16b | Present a flow diagram that gives separately the number of studies focused on harms outcomes. | | | | |  |
|  | 16c**^c^** | List the studies (full citation) that were excluded after reading the full text and provide reasons. | | | | | S4 |
| 17. Review & primary study characteristics | 17a**^c^** | Describe characteristics of each included SR in tables (such as title or author, search date, PICOS, design and number of studies included, number and age range of participants, dose/frequency, follow up period [treatment duration], review limitations, results or conclusion) and of each additional primary study. | | | | | 4, Table 1 |
|  | 17b | For each included SR report language and publication status restrictions that have been used. | | | | | - |
| 18. Overlapping | 18 | Present or/and discuss about overlapping of studies within SRs (at least one of the following): | | | | | 4, Fig 2 |
|  |  | Present measures of overlap (such as CCA). | | | | | Fig 2 |
|  |  | Provide citation matrix.**^c^** | | | | | S6 |
|  |  | Give the number of index publications or/and discuss about overlapping.**^f^** | | | | | 5, 16 |
| 19. Present assessment of methodological quality & quality of evidence | 19 | Present results in text or/and tables**^c^** of any quality assessment (see also subitems 13a-c): | | | | | S7 |
|  |  | Reporting or/and methodological quality of the included SRs. | | | | | 4, S7 |
|  |  | Inform for the quality of the individual studies that were included in the SRs (report results for sequence generation, allocation concealment, blinding, withdrawals, bias etc.) and for the additional included primary studies. | | | | | NA |
|  |  | Quality of evidence. | | | | | Table 3 |
| 20. Present meta-bias(es) | 20 | Present results of any assessment of meta-bias(es) (such as publication bias or selective reporting across studies, ROBIS assessment). | | | | | NA |
| 21. Synthesis of results | 21a | Summarize and present the main findings of the overview for benefits and harms. If a quantitative synthesis has been conducted, present each summary measure with a confidence interval, prediction interval or a credible interval and measures of heterogeneity or inconsistency. | | | | | 5, 6, 7 Table 2 S8 |
|  | 21b | Give results of any additional analyses (such as sensitivity, subgroup analyses, or meta-regression). | | | | | 5,6,7, table 2, S8 |
|  | 21c | Report results for adverse events separately for each intervention. | | | | | S8 |
| **DISCUSSION** | | | | | | |  |
| 22. Summary of evidence | 22 | Provide a concise summary of the main findings with the strength and shortcomings of evidence for each main outcome. | | | | | 15 |
| 23. Limitations | 23a | Discuss limitations of either the overview or included studies (or both) (e.g. different eligibility criteria, limitations of searching reviews, language restrictions, publication and selection bias). | | | | | 16 |
|  | 23b | Report possible limitations of the included reviews related to harms (issues of missing data and information, definitions of harms, rare adverse effects). | | | | | 16 |
| 24. Conclusions | 24a | Provide a general interpretation of the results in coherence with the review findings and present implications for practice; consider the harms equally as carefully as the benefits and in the context of other evidence. | | | | | 17 |
|  | 24b | Present implications for future research. | | | | | 17 |
| **AUTHORSHIP** | | | | | | |  |
| 25. Contributions of authors | 25 | Provide contributions of authors. | | | | | 18 |
| 26. Dual (co-)authorship | 26 | Report about dual (co-)authorship in the limitation or declarations of interest section. | | | | | 18 |
| **FUNDING** | | | | | | |  |
| 27. Funding or other support | 27a | Indicate sources of financial and other support for the OoSRs (direct funding) or for the authors (indirect funding), or report no funding. | | | | | 18 |
|  | 27b | Provide name for the overview funder and/or sponsor, or for the authors’ supporters. | | | | | 18 |
|  | 27c | Describe roles of funder(s), sponsor(s), and/or institution(s), if any, in conducted the OoSRs. | | | | | 18 |

***Abbreviations:*** *OoSRs, Overview of Systematic Reviews; SRs, Systematic Reviews; PICOS, participants, interventions, comparisons, outcomes, and study design; CCA, corrected covered area.*

^a^Applicable mainly for OoSRs that focus on adverse events. The description could be placed in methods section. ^b^Language restrictions, publication status, and years could also be reported in information sources topic—see item 7. ^c^It could also be placed in an appendix as a supplementary material. ^d^The software used for the management of the records and data could be placed in the data collection process—see item 11. ^e^The way of evaluation (e.g. instruments) can be reported in item 19. ^f^Index publication is the first occurrence of a primary publication in the included reviews. Discussion for overlapping might be placed in the discussion section.

*Modified and extended for Overviews of Systematic Reviews (OoSRs) from:* Moher D, Liberati A, Tetzlaff J, Altman DG, The PRISMA Group (2009). Preferred Reporting Items for Systematic Reviews and Meta-Analyses: The PRISMA Statement. PLoS Med 6(7): e1000097. <https://doi.org/10.1371/journal.pmed.1000097>
