## Supplementary material for "COVID-19 and pregnancy: An umbrella review of clinical presentation, vertical transmission, and maternal and perinatal outcomes": S1 to S10: S2. Outcomes of interest.docx

| Dimension to be evaluated | **Outcome** |
| --- | --- |
| Pregnancy Outcomes | -Maternal Death  -Miscarriage or Abortion  -Fetal distress  -Intrauterine growth retardation  -Placenta previa  -Placenta abruptio  -PROM  -Stillbirth  -Hypertension  -Preeclampsia  -Gestational diabetes  -C-section  -Vaginal delivery |
| Clinical Presentation in  Pregnant women | -Mechanical ventilation  -Admission to IUC  -Sepsis  -Pneumonia  -Dyspnea  -Mild respiratory symptoms  -Fever  -Headache  -Fatigue/malaise  -Myalgia  -Diarrhea  -Asymptomatic |
| Neonatal Outcomes | -Mortality  -Admission to NICU  -Mechanical ventilation  -Low birth weight  -Small for gestational age  -Low Apgar (<7)  -Preterm (<37 weeks)  -Congenital anomalies  -Neonatal birth weights |
| Clinical Presentation in Neonates | -Sepsis  -Respiratory distress syndrome  -Mild respiratory symptoms  -Fever  -Shortness of Breath  -Gastrointestinal symptoms  -Asymptomatic |
| Laboratory and radiological findings  in Pregnant Women | -Reactive C protein  -Lymphocytopenia  -Leukocytosis  -Thrombocytopenia  -Elevated ALT or AST  -Elevated D-Dimer  -Signs of Pneumonia on x-ray or CT |
| Laboratory and radiological findings  in Neonates | -Elevated SARS-CoV-2 IgM  -Elevated SARS-CoV-2 IgG  -Confirmed RT-PCR  -Radiology Pneumonia |
