## Supplementary material for "COVID-19 and pregnancy: An umbrella review of clinical presentation, vertical transmission, and maternal and perinatal outcomes": S1 to S10: S3. Search strategy.docx

**PubMed**

**Search	Query**

#53	 #20 AND #51 AND #52

#52	 (Systematic Review[sb] OR Systematic Review[tiab] OR Meta-Analysis[pt] OR Meta-Analys*[tiab] OR "Cochrane Database Syst Rev"[ta] OR Metaanalysis[tiab] OR Metanalysis[tiab] OR Overview[ti] OR (Review[ti] AND Literature[ti]) OR (MEDLINE[tiab] AND Cochrane[tiab]))

#51	 #21 OR #22 OR #23 OR #24 OR #25 OR #26 OR #27 OR #28 OR #29 OR #30 OR #31 OR #32 OR #33 OR #34 OR #35 OR #36 OR #37 OR #38 OR #39 OR #40 OR #41 OR #42 OR #43 OR #44 OR #45 OR #46 OR #47 OR #48 OR #49 OR #50

#50	 Miscarriage*[tiab]

#49	 Abortion*[tiab]

#48	 Abortion, Spontaneous[Mesh]

#47	 Partum[tiab]

#46	 Parturition*[tiab]

#45	 Childbirth*[tiab]

#44	 Parturition[Mesh]

#43	 Gestational[tiab]

#42	 Perinatal[tiab]

#41	 Perinatal Care[Mesh]

#40	 Mother*[tiab]

#39	 Mothers[Mesh]

#38	 Intrauterine[tiab]

#37	 Vertical[tiab]

#36	 Mother-to-Child[tiab]

#35	 Infectious Disease Transmission, Vertical[Mesh]

#34	 Fetomaternal[tiab]

#33	 Materno Fetal[tiab]

#32	 Maternofetal[tiab]

#31	 Fetus[tiab]

#30	 Fetal[tiab]

#29	 Fetus[Mesh]

#28	 Pregnan*[tiab]

#27	 Pregnancy Complications, Infectious[Mesh]

#26	 Pregnancy[Mesh]

#25	 Ante-Natal[tiab]

#24	 Antenatal[tiab]

#23	 Pre-Natal Care*[tiab]

#22	 Prenatal Care*[tiab]

#21	 Prenatal Care[Mesh]

#20	 #1 OR #2 OR #3 OR #4 OR #5 OR #6 OR #7 OR #8 OR #9 OR #10 OR #11 OR #12 OR #13 OR #14 OR #15 OR #16 OR #17 OR #18 OR #19

#19	 N Cov[tiab]

#18	 CV-19[tiab]

#17	 CV19*[tiab]

#16	 HCov*[tiab]

#15	 Corono Virus[tiab]

#14	 Virus Corona[tiab]

#13	 Coronovir*[tiab]

#12	 Coronavir*[tiab]

#11	 Pneumonia[tiab] AND Wuhan[tiab] AND 2019[tiab]

#10	 SARS-CoV2[tiab]

#9	 SARS-CoV-2[tiab]

#8	 2019-nCoV[tiab]

#7	 COVID19*[tiab]

#6	 COVID-19[tiab]

#5	 Corona Virus[tiab]

#4	 COVID-19[Supplementary Concept]

#3	 Severe Acute Respiratory Syndrome Coronavirus 2[Supplementary Concept]

#2	 Spike protein, SARS-CoV-2 [Supplementary Concept]

#1	 Coronavirus[Mesh]

**Cochrane Library (Wiley)**

ID	Search

#1	MeSH descriptor: [Coronavirus] explode all trees

#2	(Corona NEAR/1 Virus):ti,ab,kw

#3	COVID-19:ti,ab,kw

#4	COVID19*:ti,ab,kw

#5	"2019-nCoV":ti,ab,kw

#6	"SARS-CoV-2":ti,ab,kw

#7	SARS-CoV2:ti,ab,kw

#8	(Pneumonia AND Wuhan AND 2019):ti,ab,kw

#9	Coronavir*:ti,ab,kw

#10	Coronovir*:ti,ab,kw

#11	(Corono NEAR/1 Virus):ti,ab,kw

#12	HCov*:ti,ab,kw

#13	CV19*:ti,ab,kw

#14	CV-19:ti,ab,kw

#15	N-Cov:ti,ab,kw

#16	#1 OR #2 OR #3 OR #4 OR #5 OR #6 OR #7 OR #8 OR #9 OR #10 OR #11 OR #12 OR #13 OR #14 OR #15

#17	MeSH descriptor: [Prenatal Care] explode all trees

#18	(Prenatal NEAR/1 Care*):ti,ab,kw

#19	(Pre-Natal NEAR/1 Care*):ti,ab,kw

#20	Antenatal:ti,ab,kw

#21	Ante-Natal:ti,ab,kw

#22	MeSH descriptor: [Pregnancy] explode all trees

#23	MeSH descriptor: [Pregnancy Complications, Infectious] explode all trees

#24	Pregnan*:ti,ab,kw

#25	MeSH descriptor: [Fetus] explode all trees

#26	Fetal:ti,ab,kw

#27	Fetus:ti,ab,kw

#28	Maternofetal:ti,ab,kw

#29	(Materno NEAR/2 Fetal):ti,ab,kw

#30	Fetomaternal:ti,ab,kw

#31	MeSH descriptor: [Infectious Disease Transmission, Vertical] explode all trees

#32	"Mother-to-Child":ti,ab,kw

#33	Vertical:ti,ab,kw

#34	Intrauterine:ti,ab,kw

#35	MeSH descriptor: [Mothers] explode all trees

#36	Mother*:ti,ab,kw

#37	MeSH descriptor: [Perinatal Care] explode all trees

#38	Perinatal:ti,ab,kw

#39	Gestational:ti,ab,kw

#40	MeSH descriptor: [Parturition] explode all trees

#41	Childbirth*:ti,ab,kw

#42	Parturition*:ti,ab,kw

#43	Partum:ti,ab,kw

#44	MeSH descriptor: [Abortion, Spontaneous] explode all trees

#45	Abortion*:ti,ab,kw

#46	Miscarriage*:ti,ab,kw

#47	#17 OR #18 OR #19 OR #20 OR #21 OR #22 OR #23 OR #24 OR #25 OR #26 OR #27 OR #28 OR #29 OR #30 OR #31 OR #32 OR #33 OR #34 OR #35 OR #36 OR #37 OR #38 OR #39 OR #40 OR #41 OR #42 OR #43 OR #44 OR #45 OR #46

#48	#16 AND #47 in Cochrane Reviews

**EMBase (Elsevier)**

No. Query

#50. #48 AND #

#49. 'systematic review':ti,ab OR 'meta análisis (topic)':pt OR 'meta analysis':ti,ab OR cochrane:jt OR metaanalysis:ti,ab OR metanalysis.ti,ab. OR (medline:ab AND cochrane:ab) OR overview:ti OR (review:ti AND literature:ti)

#48. #17 AND #47

#47. #18 OR #19 OR #20 OR #21 OR #22 OR #23 OR #24 OR #25 OR #26 OR #27 OR #28 OR #29 OR #30 OR #31 OR #32 OR #33 OR #34 OR #35 OR #36 OR #37 OR #38 OR #39 OR #40 OR #41 OR #42 OR #43 OR #44 OR #45 OR #46

#46. miscarriage*:ti,ab

#45. abortion*:ti,ab

#44. 'spontaneous abortion'/exp

#43. partum:ti,ab

#42. parturition*:ti,ab

#41. childbirth*:ti,ab

#40. 'birth'/exp

#39. gestational:ti,ab

#38. perinatal:ti,ab

#37. 'perinatal care'/exp

#36. mother*:ti,ab

#35. 'mother'/exp

#34. intrauterine:ti,ab

#33. vertical:ti,ab	1

#32. 'mother-to-child':ti,ab

#31. 'vertical transmission'/exp

#30. fetomaternal:ti,ab

#29. (materno NEAR/1 fetal):ti,ab

#28. maternofetal:ti,ab

#27. fetus:ti,ab

#26. fetal:ti,ab

#25. 'fetus'/exp

#24. pregnan*:ti,ab

#23. 'pregnancy'/exp

#22. 'ante natal':ti,ab

#21. antenatal:ti,ab

#20. ('pre natal' NEAR/1 care*):ti,ab

#19. (prenatal NEAR/1 care*):ti,ab

#18. 'prenatal care'/exp

#17. #1 OR #2 OR #3 OR #4 OR #5 OR #6 OR #7 OR #8 OR #9 OR #10 OR #11 OR #12 OR #13 OR #14 OR #15 OR #16

#16. 'n cov':ti,ab

#15. 'cv 19':ti,ab

#14. cv19*:ti,ab

#13. hcov*:ti,ab

#12. (corono NEAR/1 virus):ti,ab

#11. coronovir*:ti,ab

#10. coronavir*:ti,ab

#9. pneumonia:ti,ab AND wuhan:ti,ab AND 2019:ti,ab

#8. 'sars cov2':ti,ab

#7. 'sars cov 2':ti,ab

#6. '2019 ncov':ti,ab

#5. covid19*:ti,ab

#4. 'covid 19':ti,ab

#3. (corona NEAR/1 virus):ti,ab

#2. 'coronavirus disease 2019'/exp

#1. 'coronavirinae'/exp

**LILACS**

(MH Coronavirus OR COVID-19 OR COVID19$ OR Corona OR Corono OR 2019-nCoV OR SARS-CoV-2 OR SARS-CoV2 OR Coronavir$ OR Coronovir$ OR HCov$ OR CV19$ OR CV-19 OR N-Cov) AND (MH Prenatal Care OR Prenatal-Care$ OR Pre-Natal-Care$ OR Antenatal OR Ante-Natal OR MH Pregnancy OR MH Pregnancy Complications, Infectious OR Pregnan$ OR Embarazo$ OR Gravid$ OR MH Fetus OR Feto OR Fetus OR Fetal OR Maternofetal OR Materno-Fetal OR Fetomaternal OR MH Infectious Disease Transmission, Vertical OR Mother-to-Child OR Vertical OR Intrauterin$ OR MH Mothers OR Mother$ OR Madre$ OR Maes OR Mae OR MH Perinatal Care OR Perinatal OR Gestational OR Gestacional OR MH Parturition OR Childbirth$ OR Parto OR Partum OR MH Abortion, Spontaneous OR Abortion OR Aborto OR Miscarriage$) AND (MH Metanálisis como Asunto OR PT metanalisis OR Meta-Anális$ OR Metaanális$ OR metanalis$ OR ((revisión OR Review OR Revisao) AND (sistematic$ OR Systematic)) ) [Words]

**Web of Science (SCI-EXPANDED, SSCI, A&HCI, ESCI)**

\# 51	#50 AND #49

\# 50	TS=Systematic Review OR TI=Systematic Review OR TS=Meta-Analysis OR TI=Meta-Analys* OR TI=Metaanalysis OR TI=Metanalysis OR TI=Overview OR SO=(COCHRANE DATABASE OF SYSTEMATIC REVIEWS)

\# 49	#48 AND #17

\# 48	#18 OR #19 OR #20 OR #21 OR #22 OR #23 OR #24 OR #25 OR #26 OR #27 OR #28 OR #29 OR #30 OR #31 OR #32 OR #33 OR #34 OR #35 OR #36 OR #37 OR #38 OR #39 OR #40 OR #41 OR #42 OR #43 OR #44 OR #45 OR #46 OR #47

\# 47	TI=Miscarriage*

\# 46	TI=Abortion*

\# 45	TS=Abortion, Spontaneous

\# 44	TI=Partum

\# 43	TI=Parturition*

\# 42	TI=Childbirth*

\# 41	TS=Parturition

\# 40	TI=Gestational

\# 39	TI=Perinatal

\# 38	TS=Perinatal Care

\# 37	TI=Mother*

\# 36	TS=Mothers

\# 35	TI=Intrauterine

\# 34	TI=Vertical

\# 33	TI=Mother-to-Child

\# 32	TS=Infectious Disease Transmission, Vertical

\# 31	TI=Fetomaternal

\# 30	TI=Materno Fetal

\# 29	TI=Maternofetal

\# 28	TI=Fetus

\# 27	TI=Fetal

\# 26	TS=Fetus

\# 25	TI=Pregnan*

\# 24	TS=Pregnancy Complications, Infectious

\# 23	TS=Pregnancy

\# 22	TI=Ante-Natal

\# 21	TI=Antenatal

\# 20	TI=Pre-Natal Care*

\# 19	TI=Prenatal Care*

\# 18	TS=Prenatal Care

\# 17	#1 OR #2 OR #3 OR #4 OR #5 OR #6 OR #7 OR #8 OR #9 OR #10 OR #11 OR #12 OR #13 OR #14 OR #15 OR #16

\# 16	TI=N Cov

\# 15	TI=CV-19

\# 14	TI=CV19*

\# 13	TI=HCov*

\# 12	TI=Corono Virus

\# 11	TI=Virus Corona

\# 10	TI=Coronovir*

\# 9	TI=Coronavir*

\# 8	TI=SARS-CoV2

\# 7	TI=SARS-CoV-2

\# 6	TI=2019-nCoV

\# 5	TI=COVID19*

\# 4	TI=Corona Virus

\# 3	TI=COVID-19

\# 2	TS=COVID-19

\# 1	TS=Corona Virus

**CNKI (ENG** **(E) Medicine & Public Health**)

TI=Covid-19 OR TI=2019-nCoV OR TI=SARS-CoV-2 OR TI=SARS-CoV2 OR TI=Coronavirus OR TI=Covid19) AND (TI=Prenatal OR TI=Antenatal OR TI=Perinatal OR TI=Pregnancy OR TI=Fetal OR TI=Maternofetal OR TI=Fetomaternal OR TI=Vertical OR TI=Mother OR TI=Maternal OR TI=Childbirth OR TI=Partum OR TI=Parturition OR TI=Abortion OR TI=Miscarriage)

**LOVE**

Prenatal Care* OR Pre-Natal Care* OR Antenatal OR Ante-Natal OR Pregnan* OR Fetal OR Fetus OR Maternofetal OR Materno Fetal OR Fetomaternal OR Mother-to-Child OR Vertical OR Intrauterine OR Mother* OR Perinatal OR Gestational OR Childbirth* OR Parturition* OR Partum OR Abortion* OR Miscarriage*) Filter results: Systematic Review
