## Supplementary material for "COVID-19 and pregnancy: An umbrella review of clinical presentation, vertical transmission, and maternal and perinatal outcomes": S1 to S10: S4. Excluded studies & exclusion reasons.docx

| Authors (2020) | Title of the study | Exclusion reason |
| --- | --- | --- |
| Ashary[1] | Single-Cell RNA-seq Identifies Cell Subsets in Human Placenta That Highly Expresses Factors Driving Pathogenesis of SARS-CoV-2 | Wrong outcomes |
| Azarkish[2] | Impact of COVID-19 infection on maternal and neonatal outcomes: a review of 287 pregnancies | Unmet DARE definition of SR |
| Boyadzhieva[3] | Coronavirus disease 2019 (COVID-19) during pregnancy in patients with rheumatic diseases | Unmet DARE definition of SR |
| Brown[4] | Perinatal mental health and the COVID-19 pandemic | Unmet DARE definition of SR |
| Cabero-Pérez[5] | [Infection by SARS-CoV-2 in pregnancy and possibility of transmission to neonates: A systematic revision] | Unmet DARE definition of SR |
| Campos[6] | Increasing maternal mortality associated with COVID-19 and shortage of intensive care is a serious concern in low‐resource settings | Unmet DARE definition of SR |
| Capobianco[7] | COVID-19 in pregnant women: A systematic review and meta-analysis | Unmet DARE definition of SR |
| Chamseddine[8] | Pregnancy and Neonatal Outcomes in SARS-CoV-2 Infection: a systematic review | Unmet DARE definition of SR |
| Chang[9] | Clinical characteristics and diagnostic challenges of pediatric COVID-19: A systematic review and meta-analysis | Wrong outcomes |
| Dawei[10] | Asymptomatic COVID-19 infection in late pregnancy indicated no vertical transmission | Unmet DARE definition of SR |
| Delfino[11] | SARS‐CoV‐2 possible contamination of genital area: implications for sexual and vertical transmission routes | Unmet DARE definition of SR |
| Delgado[12] | Rates of Maternal and Perinatal Mortality and Vertical Transmission in Pregnancies Complicated by Severe Acute Respiratory Syndrome Coronavirus 2 (SARS-Co-V-2) Infection: A Systematic Review | Unmet DARE definition of SR |
| Di Mascio[13] | Outcome of coronavirus spectrum infections (SARS, MERS, COVID-19) during pregnancy: a systematic review and meta-analysis | Duplicated |
| Di Nardo[14] | A literature review of 2019 novel coronavirus (SARS-CoV2) infection in neonates and children | Unmet DARE definition of SR |
| Dubey[15] | Maternal and neonatal characteristics and outcomes among COVID-19 infected women: An updated systematic review and meta-analysis | Unmet DARE definition of SR |
| Duran[16] | COVID-19 and newborn health: systematic review | Duplicated |
| Elshafeey[17] | A systematic scoping review of COVID-19 during pregnancy and childbirth | Unmet DARE definition of SR |
| Furlan[18] | A Systematic Review of Pregnancy and Coronavirus Infection: Maternal, Fetal and Neonatal Outcomes | Duplicated |
| Galang[19] | Severe Coronavirus Infections in Pregnancy: A Systematic Review | Unmet DARE definition of SR |
| Gillian[20] | Clinical update on COVID-19 in pregnancy: A review article | Unmet DARE definition of SR |
| Hoekstra[21] | Corticosteroid use in COVID-19 patients: A systematic review and meta-analysis on clinical outcomes | Wrong population |
| Ibrahim[22] | Review of published systematic reviews and meta-analyses on COVID-19 | Wrong population |
| IETSI[23] | [SARS-COV-2 mother-fetus transmission: rapid synthesis of evidence] | Unmet DARE definition of SR |
| Irani[24] | Novel coronavirus disease 2019 and perinatal outcomes | Unmet DARE definition of SR |
| Jahangir[25] | Clinical manifestations and outcomes of COVID-19 in the paediatric population: a systematic review | Unmet DARE definition of SR |
| Juan[26] | Effects of Coronavirus Disease 2019 (COVID-19) on Maternal, Perinatal and Neonatal Outcomes: a Systematic Review of 266 Pregnancies | Duplicated |
| Juan[27] | COVID-19/SARS-CoV-2 News from Preprints; Effects of Coronavirus Disease 2019 (COVID-19) on Maternal, Perinatal and Neonatal Outcomes: a Systematic Review of 266 Pregnancies | Unmet DARE definition of SR |
| Kahathuduwa[28] | Case fatality rate in COVID-19: a systematic review and meta-analysis | Wrong outcomes |
| Khalil[29] | Coronavirus - COVID-19; Findings from A. Khalil and Co-Researchers Advance Knowledge in COVID-19 Outcome of Coronavirus spectrum infections (SARS, MERS, COVID 1 -19) during pregnancy: a systematic review and meta-analysis | Unmet DARE definition of SR |
| Khalili[30] | Male Fertility and the COVID-19 Pandemic: Systematic Review of the Literature | Unmet DARE definition of SR |
| Klaritsch[31] | COVID-19 During Pregnancy and Puerperium - A Review by the Austrian Society of Gynaecology and Obstetrics (OEGGG) | Unmet DARE definition of SR |
| Lubbe[32] | Breastfeeding during the COVID-19 pandemic - a literature review for clinical practice | Unmet DARE definition of SR |
| Mark[33] | Community-Onset SARS-CoV-2 Infection in Young Infants: A Systematic Review | Unmet DARE definition of SR |
| Martins-Filho[34] | [To breastfeed or not to breastfeed? Lack of evidence on the presence of SARS-CoV-2 in breastmilk of pregnant women with COVID-19] | Duplicated |
| Mascarenhas[35] | COVID-19 and the production of knowledge regarding recommendations during pregnancy: a scoping review | Unmet DARE definition of SR |
| Melekhina[36] | Clinical characteristics of covid-19 in children of different ages. Literature review as of april 2020 | Unmet DARE definition of SR |
| Mojgan[37] | Vertical Transmission of Coronavirus Disease 19 (COVID-19) from Infected Pregnant Mothers to Neonates: A Review | Unmet DARE definition of SR |
| Mullins[38] | Coronavirus in pregnancy and delivery: rapid review | Unmet DARE definition of SR |
| Mullins[39] | COVID-19/SARS-CoV-2 News from Preprints; Coronavirus in Pregnancy and Delivery: Rapid Review and Expert Consensus (Published March 8, 2020) | Unmet DARE definition of SR |
| Mullins[40] | Coronavirus - COVID-19; Recent Findings from North Bristol NHS Trust Has Provided New Data on COVID-19 (Coronavirus in pregnancy and delivery: rapid review) | Unmet DARE definition of SR |
| Oskovi-Kaplan[41] | Coronavirus - COVID-19; New Findings from Maternity Hospital in the Area of COVID-19 Reported (The Effect of COVID-19 Pandemic and Social Restrictions on Depression Rates and Maternal Attachment in Immediate Postpartum Women: a Preliminary Study) | Unmet DARE definition of SR |
| Pastick[42] | A Systematic Review of Treatment and Outcomes of Pregnant Women With COVID-19-A Call for Clinical Trials | Unmet DARE definition of SR |
| Priputnevich[43] | The novel coronavirus SARS-COV-2 and pregnancy: Literature review | Unmet DARE definition of SR |
| Rafat[44] | Setting realistic goals for feeding infants when their mothers have suspected or confirmed COVID-19 | Unmet DARE definition of SR |
| Ramadhani[45] | Can remdesivir treat COVID-19 effectively in hospitalized pregnancies?: A literature review | Unmet DARE definition of SR |
| Rodrigues[46] | Pregnancy and breastfeeding during COVID-19 pandemic: A systematic review of published pregnancy cases | Unmet DARE definition of SR |
| Rolnik[47] | Can COVIDâ€19 in pregnancy cause preâ€eclampsia? | Unmet DARE definition of SR |
| Samantha[48] | Psychological impact of infectious disease outbreaks on pregnant women: Rapid evidence review | Wrong population |
| Sampieri[49] | [Review of new evidence about the possible vertical transmission of coronavirus disease-2019] | Unmet DARE definition of SR |
| Shorey[50] | Lessons from past epidemics and pandemics and a way forward for pregnant women, midwives and nurses during COVID-19 and beyond: A meta-synthesis | Wrong population |
| Silva[51] | Is SARS-CoV-2 Vertically Transmitted? | Unmet DARE definition of SR |
| Silva[52] | Immunological aspects of coronavirus disease during pregnancy: an integrative review | Unmet DARE definition of SR |
| Stumpfe[53] | SARS-CoV-2 Infection in Pregnancy - a Review of the Current Literature and Possible Impact on Maternal and Neonatal Outcome | Unmet DARE definition of SR |
| Tobaiqy[54] | Therapeutic Management of COVID-19 Patients: A systematic review | Wrong population |
| Torre[55] | [Recommendations and practical management of pregnant women with COVID-19: a scoping review] | Unmet DARE definition of SR |
| Vieira[56] | Repercussions of the covid-19 pandemic on the mental health of pregnant and puerperal women: a systematic review | Wrong population |
| Walker[57] | Maternal transmission of SARS-COV-2 to the neonate, and possible routes for such transmission: A systematic review and critical analysis | Unmet DARE definition of SR |
| Wingert[58] | Risk factors for severe outcomes of COVID-19: a rapid review | Wrong population |
| Yaqian[59] | Clinical and pathological characteristics of 2019 novel coronavirus disease (COVID-19): a systematic review | Unmet DARE definition of SR |
| Yeo[60] | Review of guidelines and recommendations from 17 countries highlights the challenges that clinicians face caring for neonates born to mothers with COVID-19 | Unmet DARE definition of SR |
