## Supplementary material for "COVID-19 and pregnancy: An umbrella review of clinical presentation, vertical transmission, and maternal and perinatal outcomes": S1 to S10: S7. Quality assessment of systematic reviews.docx

### S7.1 Quality assessment of systematic reviews by the 16 AMSTAR-2 items

| **Authors (2020)** | 1* | 2* | 3* | 4* | 5* | 6* | 7* | 8* | 9 RCT* | 9 NRS* | 10* | 11 RCT* | 11 NRS* | 12* | 13* | 14* | 15* | 16* | **# Non-negative** | **Overall confidence** |
| --- | --- | --- | --- | --- | --- | --- | --- | --- | --- | --- | --- | --- | --- | --- | --- | --- | --- | --- | --- | --- |
| **AbdelMassih[****1****]** | Yes | N | N | PY | N | Y | N | N | N | N | N | N MA | N MA | N MA | N | N | N | Y | 6 | Critically low |
| **Abdollahpour[****2****]** | Y | PY | N | Y | Y | Y | N | Y | NA | N | N | N MA | N MA | N MA | N | N | N MA | Y | 10 | Critically low |
| **Akhtar[****3****]** | Y | N | Y | Y | Y | Y | N | Y | NA | Y | N | N MA | N MA | N MA | N | Y | N MA | Y | 12 | Critically low |
| **Allotey[****4****]** | Y | Y | Y | Y | Y | Y | N | Y | NA | Y | N | NA | Y | Y | Y | Y | Y | Y | 14 | Low |
| **Arabi[****5****]** | Y | N | Y | PY | Y | N | N | Y | NA | N | N | Y | Y | Y | Y | N | N | N | 8 | Critically low |
| **Ashraf[****6****]** | Y | PY | Y | Y | N | N | N | Y | NA | N | N | N MA | N MA | N MA | N | N | N MA | Y | 9 | Critically low |
| **Banaei[****7****]** | Y | N | Y | Y | Y | Y | N | Y | NA | Y | N | N MA | N MA | N MA | N | N | N MA | Y | 11 | Critically low |
| **Bwire[****8****]** | Y | Y | Y | Y | Y | Y | N | Y | NA | N | N | N MA | N MA | N MA | Y | N | N MA | Y | 12 | Critically low |
| **Caparros Gonzalez[****9****]** | Y | N | Y | Y | N | N | N | N | NA | N | N | N MA | N MA | N MA | N | N | N MA | N | 6 | Critically low |
| **Centeno-Tablante[****10****]** | Y | Y | Y | Y | Y | Y | N | Y | NA | PY | N | NA | N MA | N MA | Y | Y | N MA | Y | 14 | Critically low |
| **Chi[****11****]** | Y | PY | Y | PY | Y | N | N | Y | NA | N | N | N MA | N MA | N MA | N | N | N MA | Y | 10 | Critically low |
| **de Sousa[****12****]** | Y | N | Y | Y | N | N | N | Y | NA | N | N | N MA | N MA | N MA | N | N | N MA | Y | 8 | Critically low |
| **Della Gatta[****13****]** | Y | Y | Y | PY | Y | Y | N | Y | NA | Y | N | N MA | N MA | N MA | N | N | N MA | N | 11 | Critically low |
| **Deniz[****14****]** | Y | N | Y | PY | N | N | N | Y | NA | N | N | N MA | N MA | N MA | N | N | N MA | N | 7 | Critically low |
| **Dhir[****15****]** | Y | N | N | PY | Y | Y | N | Y | NA | N | N | N MA | N | N MA | N | N | N | N | 6 | Critically low |
| **Di Mascio[****16****]** | Y | Y | Y | PY | Y | Y | N | Y | NA | N | N | N MA | Y | N | N | Y | N | Y | 10 | Critically low |
| **Diriba[****17****]** | Y | N | Y | PY | Y | Y | N | Y | NA | Y | N | N MA | Y | N | Y | Y | Y | Y | 12 | Critically low |
| **Duran[****18****]** | Y | N | N | PY | N | N | N | Y | NA | N | N | N MA | N MA | N MA | N | N | N MA | Y | 7 | Critically low |
| **Figueiro-Filho[****19****]** | Y | N | N | Y | Y | Y | N | Y | NA | N | N | N MA | N MA | N MA | N | N | N MA | Y | 9 | Critically low |
| **Furlan[****20****]** | Y | N | Y | Y | Y | Y | N | Y | NA | N | N | N MA | N MA | N MA | N | N | N MA | Y | 10 | Critically low |
| **Gajbhiye[****21****]** | Y | N | N | PY | Y | N | N | Y | NA | N | N | N MA | N MA | N MA | N | N | N MA | Y | 8 | Critically low |
| **Gao[****22****]** | Y | N | N | Y | Y | Y | N | Y | NA | N | N | Y | Y | N | N | Y | Y | Y | 9 | Critically low |
| **Goh[****23****]** | Y | N | N | PY | N | N | N | N | NA | N | N | N | N | N | N | N | N | Y | 3 | Critically low |
| **Gordon[****24****]** | Y | PY | Y | PY | Y | Y | N | Y | NA | Y | N | N MA | N MA | N MA | Y | N | N MA | Y | 13 | Critically low |
| **Han[****25****]** | Y | N | N | PY | N | N | N | PY | NA | N | N | NA | Y | N | N | N | N | Y | 7 | Critically low |
| **Hasan[****26****]** | Y | PY | N | Y | Y | N | N | Y | NA | N | N | N MA | N MA | N MA | Y | Y | N MA | Y | 11 | Critically low |
| **Hessami[****27****]** | Y | N | N | PY | N | N | N | Y | NA | N | N | N MA | N MA | N MA | N | N | N MA | Y | 7 | Critically low |
| **Huntley[****28****]** | Y | PY | Y | Y | Y | Y | N | Y | NA | Y | N | N MA | N MA | N MA | Y | Y | N MA | Y | 14 | Low |
| **Juan[****29****]** | Y | Y | N | Y | Y | Y | Y | Y | NA | Y | N | N MA | N MA | N MA | Y | Y | N MA | Y | 14 | Moderate |
| **Kasraeian[****30****]** | Y | N | N | Y | N | N | N | Y | N | N | N | Y | Y | N | N | N | Y | Y | 6 | Critically low |
| **Khalil[****31****]** | Y | Y | N | PY | Y | Y | Y | Y | NA | PY | N | NA | Y | Y | N | Y | Y | Y | 13 | Critically low |
| **Khan[****32****]** | Y | N | Y | N | N | Y | N | Y | NA | Y | N | N MA | N MA | N MA | N | N | N MA | Y | 9 | Critically low |
| **Kotlyar[****33****]** | Y | Y | N | Y | Y | N | N | Y | NA | PY | N | N | N | N | N | N | N | Y | 7 | Critically low |
| **Li[****34****]** | Y | PY | Y | Y | Y | Y | Y | PY | NA | PY | N | N MA | N MA | N MA | N | N | N MA | N | 7 | Critically low |
| **Martins[****35****]** | Y | N | N | N | N | N | N | Y | NA | N | N | N MA | N MA | N MA | N | N | N MA | N | 5 | Critically low |
| **Matar[****36****]** | Y | N | N | Y | N | N | N | Y | NA | Y | N | N MA | Y | N | N | Y | Y | N | 7 | Critically low |
| **Melo[****37****]** | Y | N | Y | PY | Y | Y | N | Y | NA | Y | N | N MA | Y | N | N | Y | N | N | 9 | Critically low |
| **Authors (2020)** | 1* | 2* | 3* | 4* | 5* | 6* | 7* | 8* | 9 RCT* | 9 NRS* | 10* | 11 RCT* | 11 NRS* | 12* | 13* | 14* | 15* | 16* | **# Non-negative** | **Overall confidence** |
| **Mirbeyk[****38****]** | Y | N | Y | Y | N | N | N | Y | NA | N | N | N MA | N MA | N MA | N | N | N MA | Y | 8 | Critically low |
| **Muhidin[****39****]** | Y | N | Y | Y | Y | Y | N | Y | NA | Y | N | N MA | N MA | N MA | N | N | N MA | Y | 11 | Critically low |
| **Mullins[****40****]** | Y | N | Y | N | N | N | N | Y | NA | N | N | N MA | N MA | N MA | N | N | N MA | Y | 7 | Critically low |
| **Mustafa[****41****]** | Y | N | N | PY | N | N | N | N | NA | N | N | N MA | N MA | N MA | N | N | N MA | Y | 6 | Critically low |
| **Panahi[****42****]** | Y | N | Y | Y | Y | N | N | Y | NA | PY | N | N MA | N MA | N MA | N | N | N MA | Y | 10 | Critically low |
| **Pettirosso[****43****]** | Y | N | Y | PY | N | N | Y | Y | NA | N | N | N MA | N MA | N MA | Y | Y | N MA | Y | 11 | Critically low |
| **Rahman[****44****]** | Y | N | Y | Y | N | N | N | Y | NA | N | N | N MA | N MA | N MA | N | N | N MA | Y | 8 | Critically low |
| **Raschetti[****45****]** | Y | PY | Y | Y | Y | Y | N | Y | NA | N | N | N MA | N MA | N MA | N | N | N MA | Y | 11 | Critically low |
| **Rodrí­guez-Blanco[****46****]** | Y | N | Y | PY | N | Y | N | Y | NA | Y | N | N MA | N MA | N MA | N | N | N MA | Y | 10 | Critically low |
| **Rostami[****47****]** | Y | N | N | Y | N | N | N | Y | NA | N | N | N MA | N MA | N MA | N | N | N MA | Y | 7 | Critically low |
| **Segars[****48****]** | Y | N | N | PY | Y | N | N | Y | NA | N | N | N MA | N MA | N MA | Y | N | N MA | N | 8 | Critically low |
| **Shi[****49****]** | Y | N | N | PY | N | N | N | Y | NA | N | N | N MA | Y | N | N | Y | N | Y | 6 | Critically low |
| **Singh[****50****]** | Y | N | N | Y | Y | Y | N | Y | NA | N | N | N MA | N MA | N MA | N | N | N MA | Y | 9 | Critically low |
| **Smith[****51****]** | Y | PY | Y | Y | Y | N | N | Y | NA | Y | N | N MA | N MA | N MA | Y | N | N MA | Y | 12 | Low |
| **Soheili[****52****]** | Y | N | Y | Y | Y | Y | N | Y | NA | Y | N | N MA | Y | N | N | N | Y | Y | 10 | Critically low |
| **Teles Abrao Trad[****53****]** | Y | N | N | PY | Y | N | N | Y | NA | N | N | N MA | N MA | N MA | N | N | N MA | Y | 8 | Critically low |
| **Thomas[****54****]** | Y | N | N | PY | N | N | N | N | NA | Y | N | N MA | N MA | N MA | N | N | N MA | Y | 7 | Critically low |
| **Trevisanuto[****55****]** | Y | N | Y | Y | Y | Y | N | Y | NA | Y | Y | N MA | N MA | N MA | N | N | N MA | Y | 12 | Critically low |
| **Trippella[****56****]** | Y | N | Y | Y | Y | Y | Y | Y | NA | Y | N | N MA | N MA | N MA | Y | Y | N MA | Y | 14 | Low |
| **Trocado[****57****]** | Y | N | Y | PY | Y | Y | N | Y | NA | Y | N | N MA | N MA | N MA | N | N | N MA | Y | 11 | Critically low |
| **Turan[****58****]** | Y | N | N | PY | Y | N | N | N | NA | N | N | N MA | N MA | N MA | N | N | N MA | Y | 7 | Critically low |
| **Uygun-Can[****59****]** | Y | N | N | PY | Y | N | N | Y | NA | Y | N | N MA | Y | N | Y | N | Y | N | 8 | Critically low |
| **Vakili[****60****]** | Y | N | N | PY | N | N | N | Y | NA | N | N | N MA | N MA | N MA | Y | N | N MA | Y | 8 | Critically low |
| **Yang N[****61****]** | Y | Y | N | PY | Y | Y | N | Y | NA | Y | N | N MA | N MA | N MA | N | N | N MA | Y | 11 | Critically low |
| **Yang Z[****62****]** | Y | PY | N | PY | Y | Y | N | Y | NA | N | N | N MA | N MA | N MA | N | N | N MA | Y | 10 | Critically low |
| **Yang Z[****63****]** | Y | N | N | Y | Y | Y | N | Y | NA | N | N | N MA | N MA | N MA | N | N | N MA | Y | 9 | Critically low |
| **Yee[****64****]** | Y | N | Y | Y | Y | Y | N | Y | NA | N | N | N | N | N | N | N | N | Y | 7 | Critically low |
| **Yoon[****65****]** | Y | N | N | Y | Y | Y | N | Y | NA | N | N | N MA | N MA | N MA | Y | Y | N MA | Y | 11 | Critically low |
| **Zaigham[****66****]** | Y | N | N | PY | Y | N | N | N | NA | Y | N | N MA | N MA | N MA | N | N | N MA | Y | 8 | Critically low |

*** The 16 AMSTAR-2 items:**

1. Did the research questions and inclusion criteria for the review include the components of PICO?
2. Did the report of the review contain an explicit statement that the review methods prior to the conduct of the review and did the report justify any significant deviations from the protocol?
3. Did the review authors explain their selection of the study designs for inclusion in the review?
4. Did the review authors use a comprehensive literature search strategy?
5. Did the review authors perform study selection in duplicate?
6. Did the review authors perform data extraction in duplicate?
7. Did the review authors provide a list of excluded studies and justify the exclusions?
8. Did the review authors describe the included studies in adequate detail?
9. Did the review authors use a satisfactory technique for assessing the risk of bias (RoB) in individual studies that were included in the review? (For RCT and NRS)
10. Did the review authors report on the sources of funding for the studies included in the review?
11. If meta-analysis was performed did the review authors use appropriate methods for statistical combination of results? (For RCT and NRS)
12. If meta-analysis was performed, did the review authors assess the potential impact of RoB in individual studies on the results of the meta-analysis or other evidence synthesis?
13. Did the review authors account for RoB in individual studies when interpreting/discussing the results of the review?
14. Did the review authors provide a satisfactory explanation for, and discussion of, any heterogeneity observed in the results of the review?
15. If they performed quantitative synthesis did the review authors carry out an adequate investigation of publication bias (small study bias) and discuss its likely impact on the results of the review?
16. Did the review authors report any potential sources of conflict of interest, including any funding they received for conducting the review?

*If the answers to all signaling questions for a domain are ‘‘yes’’ or ‘‘probably yes,’’ then level of concern can be judged as low. If any signaling question is answered ‘‘no’’ or ‘‘probably no,’’ potential for concern about bias exists.

### S7.2 Quality assessment of systematic reviews by ROBIS

| **Authors (2020)** | **Phase 2: concerns with the review process** | | | | **Phase 3*** |
| --- | --- | --- | --- | --- | --- |
|  | **1. Study eligibility criteria** | **2. Identification and selection of studies** | **3. Data collection and study appraisal** | **4. Synthesis and findings** | **Risk of bias in the review** |
| **Allotey[****4****]** | **☺️** | **☺️** | **☺️** | **☺️** | **☺️ Low** |
| **Centeno-Tablante[****10****]** | **☺️** | **☺️** | **☺️** | **?** | **? Unclear** |
| **Figueiro-Filho[****19****]** | **☺️** | **?** | **☹** | **?** | **☹ High** |
| **Juan[****29****]** | **☹** | **☺️** | **☺️** | **?** | **☹ High** |

*If the answers to all signaling questions for a domain are ‘‘yes’’ or ‘‘probably yes,’’ then level of concern can be judged as low. If any signaling question is answered ‘‘no’’ or ‘‘probably no,’’ potential for concern about bias exists.
