## Supplementary material for "COVID-19 and pregnancy: An umbrella review of clinical presentation, vertical transmission, and maternal and perinatal outcomes": S1 to S10: S8. Systematic review level numerical data by research question.docx

### ***Table 1: Clinical presentation in SARS-COV2 pregnant women***

| Authors (2020) | N pregnant women | Asympto-matic | Headache | Myalgia | Fatigue/  malaise | Fever | Diarrhea | Mild respiratory symptoms | Dyspnea/  Shortness of breath | Pneumonia | Sepsis | Mechanical  ventilation/  IUC | Death |
| --- | --- | --- | --- | --- | --- | --- | --- | --- | --- | --- | --- | --- | --- |
| **AbdelMassih[****1****]** | 1787 | 219/1787 (12%) | 60/1787 (3.3%) | 56/1787 (3%) | NA | 951/1787 (53%) | 78/1787 (4.4%) | 934/1787 (52.2%) | 417/1787 (23.3%) | NA | NA | 130/1787 (7%) | NA |
| **Akhtar[****2****]** | 156 | NA | NA | 17/156 (11%) | 13% | 82/156 (53%) | NA | 50/156 (32%) | 12/156 (8%) | NA | NA | 17/156 (11%) | NA |
| **Allotey[****3****]** | depends on each outcome | 115/2380  (4.8%) | NA | 1411/6078  (23.21%) | NA | 2733/8328  (32.81%) | 659/7525  (8.74%) | 3432/8317 (41.26%) | 1928/8159(23.63%) | 729/2577  (28.88%) | 7/737 (0.94%) | 155/10713  (1.44%) | 73/11 580 (0.63%) |
| **Arabi[****4****]** | 50 | NA | NA | 5/50 (1%) | 0/50 (0%) | 38/50 (77%) | 0/50 (0%) | 10/50 (20%) | 5/50 (10%) | NA | NA | NA | 0/50 (0%) |
| **Ashraf[****5****]** | 90 | NA | 3/90 (3.3%) | 6/90 (6.6%) | 11/90  (12.2%) | 47/90 (52.2%) | 1/90 (1.1%) | 34/90 (37.7%) | 12/90  (13.3%) | 59/90 (65%) | 1/90 (1.1%) | 3/90 (3.3%) | 1/90 (1.1%) |
| **de Sousa[****6****]** | 755 | 83/689 (12%) | NA | 51/689 (7.4%) | 69/689 (10%) | 363/689 (52.7%) | 29/689 (4.2%) | NA | 78/689 (11.4%) | NA | NA | NA | NA |
| **Della Gatta[****7****]** | 51 | 13 (31%) | NA | 3/35 (8.6%) | 3/35 (8.6%) | 17/35 (48%) | 2/35 (5.7%) | 16/35 (46%) | 4/35 (11.4%) | NA | NA | NA | NA |
| **Diriba[****8****]** | depends on each outcome | NA | 88/674  (13.05%) | 61/293  (20.8%) | 192/634  (20.34%) | 575/870 (66%) | NA | 458/869  (52.7%) | 49/303  (16%) | 368/517(71.2%) | NA | NA | 15/557  (2.69%) |
| **Figueiro-Filho[****9****]** | depends on each outcome | 814/6598 (6.2%) | 1409/3474 (40.69%) | 1861/4290 (43.49%) | 55/680 (8.1%) | 2050/5138 (40%) | 597/5154 (11.6%) | 2667/5154 (51.75%) | 1303/5154 (51.75%) | NA | NA | 170/1243 (13.7%) | NA |
| **Furlan[****10****]** | 399 | NA | NA | 8/19 (42.1%) | NA | 87/157 (55.4%) | NA | 46/119 (38.6%) | 7/32  (21.8%) | NA | NA | 6/35 (17.1%) | 2/284 (0.7%) |
| **Gajbhiye[****11****]** | 441 | NA | NA | 70/369 (18.9) | NA | 206/369 (55.8) | NA | 159/369( 43%) | NA | 30/369 (8.1%) | NA | 31/369 (8.4%) | 4/369 (1%) |
| **Gao[****12****]** | 236 | NA | NA | NA | NA | 120/236 (51%) | NA | 73/236 (31%) | NA | NA | NA | NA | NA |
| **Han[****13****]** | depends on each outcome | NA | NA | 83/678 (12.68%) | NA | 568/954 (59.54%) | 37/787 (4.7%) | 506/952 (53.15%) | 241/885  (27.23%) | NA | NA | 36/221  (16.28%) | 13/1005(1.29%) |
| **Huntley[****14****]** | 538 | NA | 15/161 (9.3%) | 26/156 (16.7%) | 52/337 (15.4%) | 238/496 (48%) | NA | 211/463 (45.6%) | 274/364 (75.2%) | NA | NA | 8/263 | 0/348 (0%) |
| **Juan[****15****]** | 324 | NA | NA | 30/315(9.5%) | 29/315 (9%) | 154/315 (48.8%) | 14/315 (4.4%) | 117/315 (37%) | 46/315 (14.6%) | 190/198 (90.5%) | NA | 12/188 | 9/315 (285) |
| **Kasraeian[****16****]** | 87 | 28/87 (30%) | NA | NA | NA | 37/87 (86%) | NA | 50/87 (68%) | NA | 87/87 (100%) | NA | NA | 0/87 (0%) |
| **Khalil[****17****]** | 2567 | 253/1205 (14.5%) | 92/640 (15.0%) | 104/640 (18.9%) | 101/545 (18.5%) | 1292/1987 (63.3%) | 126/1621 (7.4%) | 391/1987 (71.4%) | 789/1941 (34.4%) | NA | NA | 92/1680 (3.4%) | 43/2468 (0.9%) |
| **Khan[****18****]** | 101 | NA | 3/101 (3%) | 7/101 (7.1%) | 15/101 (15.2%) | 66/101 (66.7%) | 4/101 (4%) | 39/101 (39.4%) | 14/101 (14.1%) | NA | NA | NA | NA |
| **Li[****19****]** | 19 | NA | NA | NA | NA | 19/19 (100%) | NA | 15/19 (78.94%) | NA | NA | NA | 5/19 (26.31%) | 3/19 (15.78%) |
| **Matar[****20****]** | 136 | NA | NA | NA | NA | 75/136 (63%) | 21/136 (15.6%) | 39/130 (37%) | 21/136 (15.7%) | NA | NA | NA | 1/136 (11.1%) |
| **Mirbeyk[****21****]** | 364 | 25/364 (7%) | NA | NA | NA | 227/364 (62.4%) | NA | 165/364 (45.3%) | NA | 22/364 (6%) | NA | NA | 2/364 (0.55%) |
| **Mullins[****22****]** | 19 | 3/19 (16%) | NA | NA | NA | NA | NA | NA | NA | NA | NA | 1/19 (5%) | NA |
| **Pettirosso[****23****]*** | 3830 | 178/510 (34.9%) | 89/198 (45%) | 137/350 (39%) | 72/317 (22.7%) | 458/989 (46.3%) | 15/295 (5%) | NA | 117/474 (24.6%) | NA | NA | NA | NA |
| **Rahman[****24****]** | 102 | NA | NA | 15/64 (23,4%) | NA | 55/64 (85.9%) | 7/79  (8.9%) | 26/79 (32.9%) | 11/79 (13.9%) | 53/70 (75.7%) | NA | 3/70 (4,3%) | 0/79  (0%) |
| **Smith[****25****]** | 92 | 30/92 (32.6%) | NA | NA | 25/82 (30.49%) | 57/92 (61.9%) | NA | 35/92 (38.4%) | 10/83 (12.1%) | 78/79 (98.7%) | NA | 1/92 (1.1%) | 2/92 (2.2%) |
| **Soheili[****26****]** | 177 | NA | NA | 10/58 (18%) | 8/58 (13%) | 71/126 (56%) | 6/65 (9%) | 33/110 (30%) | 5/176 (3%) | NA | NA | NA | NA |
| **Teles Abrao Trad[****27****]** | 139 | 29/139 (20%) | NA | NA | NA | 80/139 (57.6%) | NA | 44/139 (31.7%) | 19/139 (13.7%) | NA | NA | 5/155  (3.2%) | NA |
| **Thomas[****28****]** | 157 | 26/63 (41%) | NA | 7/93  (7.52%) | 6/93  (6.45%) | 67/93  (72%) | 6/93  (6.45%) | 27/93  (29%) | 6/93 (6.45%) | NA | NA | NA | 1/93 (1%) |
| **Trippella[****29****]** | 275 | 22/269 (8%) | NA | 37/269  (13.8%) | 28/275 (10.2%) | 155/275  (56.3%) | 9/275  (3.27%) | 98/275  (35.6%) | 9/275  (3.3%) | 162/275 (59%) | NA | NA | 1/275  (0.36%) |
| **Trocado[****30****]** | 95 | NA | NA | 6/95 (6%) | 10/95 (11%) | 52/95 (55%) | 4/95 (4%) | 36/95 (38%) | 7/95(8%) | NA | NA | NA | NA |
| **Turan[****31****]** | 637 | 68/637 (10.7%) | NA | 79/515 (15.3%) | NA | 273/515 (53%) | 18/515 (3.5%) | 224/515 (43.5%) | 64/515 (12.4%) | NA | NA | 51/61 (83.6%) | 1.6% (10/637) |
| **Uygun-Can[****32****]** | 181 | NA | NA | NA | NA | 69/181 (38%) | NA | 40/181 (22%) | 6/181 (3.3%). | NA | NA | NA | NA |
| **Yang Z[****33****]** | 114 | NA | NA | 18/114 (16.3%) | 26/114 (22.5%) | 100/114 (87.5%) | 10/114  (8.8%) | 61/114 (53.8%) | 13/114 (11.3%) | NA | NA | NA | NA |
| **Yee[****34****]** | 9032 | NA | NA | 1354/8372 (16.2%) | 60/127 (54.5%) | 1395/8571 (27.6%) | 505/8310 (6.5%) | 2018/8560 (50.1%) | 1130/8560 (20.7%) | NA | NA | NA | NA |
| **Yoon[****35****]** | 223 | NA | NA | 12/56  (21.4%) | NA | 85/201 (42.4%) | NA | 64/201  (31.8%) | 16/142 (42.4%) | NA | NA | NA | NA |
| **Zaigham[****36****]** | 108 | NA | NA | 11/108  (10.2%) | 14/108  (12.9%) | 63/92 (68%) | 7/108  (6.5%) | 37/108 (40%) | 13/108 (14%) | NA | NA | NA | 0/108 (0%) |

*Information calculated by the authors

### ***Table 2: Laboratory findings in SARS-COV2 pregnant women***

| Author | N pregnant women | Increased reactive c protein | Lymphocytopenia | Leukocytosis  / neutrophilia | Trombocyto-penia | Elevated ALT or AST | Elevated d-dimer | Radiology pneumonia | Ground-glass opacities in CT | Infiltrated shadows in CT | Signs of pneumonia XR or CT |
| --- | --- | --- | --- | --- | --- | --- | --- | --- | --- | --- | --- |
| **Allotey[****3****]** | depends on each outcome | 174/426  (40.84%) | 262/780(33.59%) | 50/251  (19.92%) | 36/428  (8.41%) | 51/491  (10.39%) | NA | NA | 246/387  (63.56%) | NA | 599/1968  (30.43%) |
| **Ashraf[****5****]** | 90 | 26/32 (81.2%) | 18/28 (64.3%) | 11/24 (45.8%) | 44/46 (95.7%) | 8/25 (32%) | NA | 59/60 (99%) | 59/60 (99%) | 59/60 (99%) | 59/60 (99%) |
| **deSousa[****6****]** | 755 | 260/475 (54.7%) | 257/475 (54.1%) | Leukocytosis 168/475 (35.4%)  Neutrophilia 40/475 (8.4%) | NA | (ALT) 39/475 (8.2%)  (AST) 45/475 (9.5%) | NA | NA | NA | NA | NA |
| **Diriba[****8****]** | depends on each outcome | 80/143  (55.94%) | 92/146  (63%) | 27/95  (28%) | NA | 9/48  (18.75%) | NA | NA | 50/76  (66.6%) | 11/19  (57.89%) | NA |
| **Figueiro-Filho[****9****]** | depends on each outcome | 162/592 (27.59%) | 146/182 (80.2%) | NA | NA | 48/427 (27.49%) | 86/385 (22.39%) | NA | NA | NA | NA |
| **Furlan[****10****]** | 399 | NA | NA | NA | 6/35  (17.14%) | 46/119 (38.6%) | 87/157  (55.4%) | 7/32  (21.8%) | NA | NA | 8/19 (42.1%) |
| **Gao[****12****]** | 236 | NA | 116/236 (49%) | NA | NA | NA | NA | NA | NA | NA | 168/236 (7.1%) |
| **Han[****13****]** | depends on each outcome | NA | 137/383 (35.8%) | NA | NA | 14/71 (19.7%) | NA | NA | NA | NA | 420/866 (48.5%) |
| **Huntley[****14****]** | 538 | NA | 110/232 (47.4%) | NA | NA | NA | NA | NA | NA | NA | NA |
| **Juan[****15****]** | 324 | 104/213  (48.8%) | 99/213(46%) | NA | NA | 12/54(22%) | NA | NA | NA | NA | 190/198(96%) |
| **Kasraeian[****16****]** | 87 | NA | 24/87 (70%) | NA | NA | NA | NA | NA | NA | NA | NA |
| **Khalil[****17****]** | 2567 | 144/351 (54.0%;) | 143/444 (34.2%) | NA | 7/259 | 48/318 (16.0%) | 77/91  (2.5%) | NA | NA | NA | NA |
| **Li[****19****]** | 19 | NA | 7/18 (38.8%) | NA | 7/18 (38.8%) | NA | NA | NA | NA | NA | 19/19 (100%) |
| **Matar[****20****]** | 136 | 43/76 (57%) | 36/72; (50%) | NA | NA | (ALT) 10/44 (22.3%)  (AST) 11/51 (23.3%) | NA | NA | 71/87 (81.7%) | 5/11 (42.5%) | NA |
| **Muhidin[****22****]** | 89 | 29/30 (96%) | 19 /30  (63%) | NA | NA | 3/30 (10%) | NA | NA | NA | NA | NA |
| **Shi[****37****]** | 173 | 104/151 (69%) | 97/164 (59%) | 37/46 (81%) | NA | NA | 89/109 (82%) | NA | NA | NA | NA |
| **Smith[****25****]** | 92 | NA | 66.7% (46/69) | NA | NA | NA | NA | NA | NA | NA | NA |
| **Soheili[****26****]** | 177 | 30/51 (58%) | 46/126 (37%) | NA | NA | NA | NA | NA | NA | NA | NA |
| **TelesAbraoTrad[****27****]** | 139 | NA | 32/90 (35.6%) | 8/90 (8.8%) | NA | NA | NA | NA | NA | NA | 82/155 (53.5% ) |
| **Trippella[****29****]** | 275 | 52/108 (48.1%) | 31/108  (7%) | NA | NA | 9/108  (26.85%) | NA | 48/48 | NA | 162/171  (95%) | NA |
| **Turan[****31****]** | 637 | 275  (72.2%) | 179  (47.0%) | 53 (13.9%) | NA | NA | 94  (24.7%) | NA | NA | NA | 196 (88.7%) |
| **Uygun-Can[****32****]** | 181 | NA | NA | NA | NA | NA | NA | NA | NA | 177/181 (97.9%) | NA |
| **Vakili[****38****]** | 54 | 36/54 (66%) | 30/54 (55%) | 41/54 (77%) | NA | 18/54 (33%) | NA | NA | NA | NA | NA |
| **Yee[****34****]** | 9032 | 86/180 (47.7%) | 108/258 (42.6%) | NA | NA | NA | NA | NA | NA | NA | NA |
| **Yoon[****35****]** | 223 | 65/103  (63.1%) | 52/120  (43.3%) | 28/89  (31.5%) | NA | NA | NA | NA | NA | NA | 185/200  (92.5%) |
| **Zaigham[****36****]** | 108 | 45/64  (70%) | 40/68  (58.8%) | NA | NA | NA | NA | NA | NA | NA | NA |

### ***Table 3: Maternal outcomes in SARS-COV2 pregnant women***

| Authors (2020) | N pregnant women | Vaginal delivery | C-section | Gestational diabetes | Preeclampsia | Hypertension | Stillbirth | PROM | Placenta previa | Miscarriage or abortion | Placenta abruptio | Intrauterine growth retardation. | Fetal distress |
| --- | --- | --- | --- | --- | --- | --- | --- | --- | --- | --- | --- | --- | --- |
| AbdelMassih[1] | 1787 | NA | NA | NA | NA | NA | NA | NA | NA | NA | NA | NA | 3/71 (7%) |
| Abdollahpour[39] | NA | NA | NA | NA | NA | NA | NA | NA | NA | NA | NA | NA | NA |
| Akhtar[2] | 156 | 19/108 (1.7%) | 66/108 (61.1%) | NA | NA | NA | NA | 86/108 (8%) | NA | NA | NA | NA | 1/108 (14%) |
| Allotey[3] | 13118 | NA | 184/491 (37.5%) | NA | NA | NA | 3/427 (0.7%) | NA | NA | NA | NA | NA | 25/293 (8.5%) |
| Arabi[4] | 50 | NA | 30/50 (61%) | NA | 0/50 (0%) | NA | NA | NA | NA | NA | NA | NA | 1/50 (0.1%) |
| Ashraf[5] | 90 | 9/90 (10%) | 81/90 (90%) | 4/90 (4.4%) | 2/90 (2.22%) | 2/90 (2.22%) | 1/90 (1.1%) | 6/90 (6.7%) | NA | NA | NA | NA | 15/86 (17,4%) |
| Banaei[40] | 123 | 17/123 (13.8%) | 99/123 (80.55) | NA | NA | NA | 1/123 (0.8%) | NA | NA | NA | NA | NA | NA |
| Chi[41] | NA | NA | NA | NA | NA | NA | NA | NA | NA | NA | NA | 10/105 (0.9%) | NA |
| Della Gatta[7] | 51 | 2/48 (4.2%) | 46/48 (95.8%) | NA | 1/34 (2.9%) | 1/48 (2.1%) | 1/34 (2.9%) | 9/34 (26.5%) | 1/34 (2.9%) | NA | 1/34 (2.9%) | NA | NA |
| Deniz[42] | 714 | 105/714 (14%) | 609/714 (85.2%) | NA | NA | NA | NA | 44/714 (6.1%) | NA | NA | NA | NA | NA |
| Dhir[43] | 1125 | NA | 742/1125 (66%) | NA | NA | NA | NA | NA | NA | NA | NA | NA | NA |
| Di Mascio[44] | 41 | NA | 38/41 (91%) | NA | 1/10 (10%) | NA | 1/42 (2.4%) | 5/31 (18.8%) | NA | NA | NA | NA | 12 /30 (40%) |
| Diriba[8] | 1316 | 198/692 (28.6%) | 440/772 (57%) | NA | 10/169 (0.6%) | NA | NA | 17/183 (9.3%) | NA | 9/62 (14.5%) | NA | 3/108 (2.7%) | 14/56 (25%) |
| Figueiro-Filho[9] | depends on each outcome | 349/1119 (31.2%) | 761/1119 (68%) | NA | NA | NA | NA | NA | NA | NA | NA | NA | NA |
| Furlan[10] | 399 | NA | NA | NA | NA | NA | 1/188 (0.5%) | NA | NA | 1/188 (0.5%) | NA | NA | NA |
| Gajbhiye[11] | 441 | NA | NA | 33/369 (8.9%) | 37/369 (10%) | NA | 4/369 (1.1%) | 33/369 (8.9%) | NA | NA | NA | NA | 31/369 (8%) |
| Gao[12] | 236 | NA | 153/236 ( 65%) | NA | NA | NA | NA | NA | NA | NA | NA | NA | 68/236 (29%) |
| Han[13] | depends on each outcome | 283/733 (38.6%) | 135/586 (23%) | NA | 24/381(6.3%) | NA | 8/663 (1.2%) | 32/176 (18.1%) | NA | 20/743 (2.7%) | NA | NA | NA |
| Huntley[14] | 538 | NA | 332/392 (85%) | NA | NA | NA | 0/274 (0.0%) | NA | NA | NA | NA | NA | NA |
| Juan[15] | 324 | 51/313 (16,3%) | 186/313 (59.4%) | 20/233 (8.6%) | 6/193 (3.1%) | 14/194 (7.2%) | 4/313 (1.3%) | NA | 1/42(2.38%) | 4/313 (1.3%) | NA | NA | NA |
| Kasraeian[16] | 87 | 1/69 (1%) | 64/69 (92%) | NA | NA | NA | 2/87 (0,2%) | 4/31(13.9%) | NA | NA | NA | NA | 10/31 (0,3%) |
| Khalil[17] | 2567 | NA | 390/746 (48.3%) | NA | NA | NA | 12/1362 (0.9%) | NA | NA | NA | NA | NA | (15/238 (5.3%) |
| Khan[18] | 101 | NA | 50/60 (83.9%) | NA | NA | NA | 0/60 (0%) | NA | NA | NA | NA | NA | NA |
| Li[19] | 19 | NA | 9/12(75%) | NA | NA | NA | NA | NA | NA | 4/10(4%) | NA | 3/13 (23%) | NA |
| Martins[45] | 24 | NA | 22/24 (91.7%) | NA | NA | NA | NA | NA | NA | NA | NA | NA | NA |
| Matar[20] | 136 | NA | 72/ 94 (76.3%) | NA | NA | NA | NA | NA | NA | NA | NA | NA | 3/94(3,2%) |
| Melo[46] | 520 | NA | 366/520 (70.4%) | NA | NA | NA | NA | NA | NA | NA | NA | NA | NA |
| Mirbeyk[21] | 364 | NA | 257/364 (86%) | NA | NA | NA | NA | NA | NA | NA | NA | NA | NA |
| Muhidin[47] | 89 | 5/86 (5.8%) | 79/86 (91.8%) | NA | 1/79 (1.3%) | NA | 2/79 (2.5%) | NA | NA | NA | 6/89 (43.2%) | NA | 15/89 (16.8%) |
| Mullins[22] | 19 | 2/19 (10.5%) | 17/19 (89%) | NA | NA | NA | NA | 8/19 (42%) | NA | NA | NA | NA | NA |
| Pettirosso[23] | 3830 | NA | NA | NA | NA | NA | 1/13(8%) | 17/160 (10.6%) | NA | 5/159 (3.14%) | NA | NA | NA |
| Rodrí­guez-Blanco[48] | NA | 8/73 (11,0%) | 65/73 (89,0%) | NA | NA | 6/64 (8.1%) | NA | 9/74 (12,1%) | NA | NA | NA | 0/74 (0,0%) | NA |
| Segars[49] | 162 | NA | NA | NA | NA | NA | NA | NA | NA | 2/162 (1.2%) | NA | 9/162 (5.5%) | 8/60 (13,3%) |
| Smith[25] | 92 | 3/50 (6%) | 40/50 (80%) | NA | NA | NA | NA | NA | NA | NA | NA | NA | 11 / 18 (61.1%) |
| Soheili[26] | 177 | 25/178 (14%) | 152/177 (86%) | NA | 46/177 (26%) | NA | 3/177 (2%) | NA | NA | NA | NA | NA | 10/65 (15%) |
| Teles Abrao Trad[27] | 139 | 9/116 (7.8%) | 107/116 (92.2%) | 10/138 (0.7%) | 4/116 (3.4%) | 5/138(3.6%) | NA | 10/116 (8.6%) | NA | NA | NA | NA | 19 / 18 (16.1%) |
| Thomas[28] | 157 | 42/157 (26.7%) | 115/157 (73.2%) | 13/44 (29%) | 6/44 (13.6%) | 5/44 (11.4%) | NA | 7/44 (15.9%) | NA | NA | 1/44 (2.3%) | NA | 6/44 (14%) |
| Trippella[29] | 275 | 60/275 (21.8%) | 179/275 (65%) | 10/275 (3.6%) | 5/275 (1.8%) | 10/275 (3.6%) | 2/275 (0.7%) | 3/275 (1%) | NA | NA | NA | NA | NA |
| Trocado[30] | 95 | 3/50 (6%) | 47/ 50 (94%) | 3/95 (3%) | 1/95 (1%) | 2/95(2%) | NA | 5/95 (5%) | 2/95 (2%) | NA | NA | NA | 7/51 (14%) |
| Turan[31] | 637 | 76/ 479 (15.9%) | 403/479 (84.1%) | NA | NA | NA | 7/500 (1.4%) | NA | NA | 7/86 (8.1%) | NA | NA | 28/60 (46.7) |
| Yang Z[33] | 114 | 9/114 (22%) | 89/114 (78%) | NA | NA | NA | 1/114 (0,9%) | NA | NA | NA | NA | NA | 8/75 (10.7%) |
| Yee[34] | 9032 | NA | NA | NA | NA | NA | NA | (2/63) 2.4% | NA | NA | NA | NA | 10/63 (15,1%) |
| Yoon[35] | 223 | NA | NA | NA | NA | NA | 2/201(1%) | 16/126(12.7%) | NA | NA | NA | NA | NA |
| Zaigham[36] | 108 | 7/85 (8.3%) | 78/85 (91.7%) | NA | NA | NA | 1/87 (1.1%) | NA | NA | NA | NA | NA | NA |

### ***Table 4: Clinical presentation in neonates born to SARS COV2 mothers***

| Author | N neonates | Asymptomatic | Reparatory distress syndrome | Sepsis | Mild respiratory symptoms | Gastrointestinal symptoms | Fever | Shortness of breath | Radiology pneumonia | SARS Cov2  (RT-PCR) | Elevated SARSCov2 IgM | Elevated SARSCov2 IgG |
| --- | --- | --- | --- | --- | --- | --- | --- | --- | --- | --- | --- | --- |
| AbdelMassih[1] | 71 | NA | NA | NA | NA | NA | 2/71 (4%) | NA | NA | NA | NA | NA |
| Akhtar[2] | 108 | NA | NA | NA | NA | 4/108 (4%) | 3/108 (3%) | 6/108 (6%) | NA | NA | NA | NA |
| Allotey[3] | 2557 | NA | NA | 2/51 (4%) | NA | NA | NA | NA | NA | NA | NA | NA |
| Ashraf[5] | 86 | 19/86 (22.1%) | 2/86 (2.32%) | NA | NA | NA | NA | NA | NA | NA | NA | NA |
| Banaei[40] | 123 | NA | 1/123 (0.8%) | NA | 1/123 (0.8%) | 2/123 (1.6%) | 3/123 (2.4%) | 1/123 (0.8%) | 4/123 (3.2%) | 5/123 (4%) | NA | NA |
| Caparros Gonzalez[50] | 10 | NA | NA | NA | NA | NA | 2/10 (20%) | 6/10 (60%) | NA | NA | NA | NA |
| Chi[41] | 105 | NA | NA | NA | NA | NA | NA | NA | NA | 5/91 (5.5%) | 3/105 (2.8%) | 3/105 (2.8%) |
| de Sousa[6] | 598 | NA | NA | NA | NA | NA | NA | NA | NA | 9/493 (2%) | 3/493 (0.6%) | 3/493 (0.6%) |
| Dhir[43] | 1141 | 13/45 (22.4%) | NA | NA | NA | 5/45 (8.6%) | 9/45 (15.5%) | 4/45 (8.8%) | NA | 39/1105 (3.9%) | NA | NA |
| Duran[51] | 222 | NA | 4/222 (8.8%) | NA | 6/222 (2.7%) | NA | 16 /222 (7.2%) | 1/222 (0.4%) | 7/71(9,8%) | NA | NA | NA |
| Figueiro-Filho [9] | Depends of each outcome | NA | 28/576 (4.9%) | 1/241 (0.4%) | NA | NA | NA | NA | NA | 18/1116 (1.6%) | NA | NA |
| Gajbhiye [11] | 391 | NA | 31 /369 (8%) | NA | NA | NA | NA | NA | 30 /369 (8%) | 17/313 (5%) | 4/8 (50%) | 7/8 (87,5%) |
| Gordon[52] | 10 | NA | NA | NA | NA | 1/10 (1%) | NA | NA | NA | 10/10 (100%) | NA | NA |
| Juan[15] | 221 | NA | NA | NA | 2 /79 (2.5%) | NA | 0 /37 (0%) | 0 /37 (0%) | 0 /37 (0%) | NA | NA | NA |
| Mirbeyk[21] | 302 | NA | NA | NA | NA | NA | NA | NA | NA | 13 /252 (5%) | NA | NA |
| Muhidin[47] | 89 | NA | NA | NA | NA | 5/89 (5,6%) | 3/89 (3,4%) | 6/89 (6,7%) | NA | 0/89 (0%) | NA | NA |
| Mullins[22] | 20 | 3 /20 (16%) | NA | NA | NA | NA | NA | NA | NA | NA | NA | NA |
| Smith[25] | 37 | 11/23 (47%) | NA | NA | NA | NA | NA | NA | NA | NA | NA | NA |
| Teles Abrao Trad[27] | 118 | NA | 10 /118 (8.5%) | NA | NA | NA | NA | NA | NA | NA | NA | NA |
| Thomas[28] | 160 | NA | NA | NA | NA | 1/13 (1%) | NA | 9/13 (5%) | NA | NA | NA | NA |
| Trevisanuto[53] | 44* | 12/38 (31.5%) | NA | NA | 7/34 (20%) | 9/34 (26%) | 17/34 (50%) | 3/34 (9%) | 15/21 (71%) | 41/44 (93%) | 3/44 (7%) | NA |
| Trippella[29] | 248 | NA | 21 /189 (11.1%) | 1/189 (0.6%) | 4/160 (2.5%) | 13 /160 (8.1%) | 13 /160 (8.1%) | NA | 6/70 (8.6%) | 7/77 (44%) | NA | NA |
| Trocado[30] | NA | NA | NA | NA | NA | NA | NA | NA | NA | 1/48 (2%) | NA | NA |
| Turan[31] | 479 | NA | 50 /479 (10%) | NA | NA | NA | NA | NA | NA | 8 /479 (1.7%) | NA | NA |
| Yang N[54] | 84 | NA | NA | NA | NA | NA | NA | NA | NA | NA | 4/84 (4.8%) | 7/84(8.3%) |
| Yang Z[55] | 83 | NA | NA | NA | NA | NA | NA | NA | NA | 5 / 154 (1.8%) | NA | NA |
| Yee[34] | 201 | 68 /73 (93.2%) | 8 /125 (6.4%) | NA | NA | 9 /125 (7.2%) | 5 /125 (4%) | 12 /125 (9.6%) | 18 /68 (26.5%) | 4 /167 (2.4%) | 3 /17 (17.6) | 6 /17 (35.3) |
| Yoon | NA | NA | NA | NA | NA | NA | NA | NA | NA | 1 /75 (1.3%) | NA | NA |

### ***Table 5: Neonatal outcomes in neonates born to SARS COV2 pregnant women***

| Author | N neonates | Neonatal birth weigth | Low birth weight (rate) | Small for Gestational age | Low Apgar (<7) | Preterm (<37 wks) | Congenital anomaly | Admission to NICU | Mechanical Ventilation | Mortality |
| --- | --- | --- | --- | --- | --- | --- | --- | --- | --- | --- |
| Akhtar[2] | 108 | NA | NA | NA | NA | 27/108 (25%) | NA | NA | NA | 10/108 (9.2%) |
| Allotey[3] | 2557 | NA | NA | NA | NA | 318/1872 (17%) | NA | 368/1348 (25%) | NA | 6/ 1728 (0.34%) |
| Arabi[4] | 50 | 3200 g mean | NA | NA | NA | 10/50 (20%) | NA | NA | NA | 0/50 (0%) |
| Ashraf[5] | 86 | 1520 - 3820 (range) | NA | 2/86 (2.3%) | NA | NA | NA | NA | NA | 1/86 (1.2%) |
| Banaei[40] | 123 | 1580 - 4000 g (range) | NA | 2/123 (1.6%) | 1/123 (0.8%) | 30/123 (24.4%) | NA | 3/123 (2.4%) | NA | 1/123 (0.8) |
| Caparros Gonzalez *[50] | 65 | NA | NA | 2/10 (20%) | NA | 6/10 (60%) | NA | NA | NA | 1/65 (1.53%) |
| Chi[41] | 105 | NA | NA | 10 (11.2%) | NA | 25(23.8%) | NA | NA | NA | 1/105 (0.9%) |
| Della Gatta[7] | 48 | NA | NA | NA | NA | 1/48 (2%) | NA | 1/48 (2%) | NA | 1/48 (2%) |
| Deniz[42] | 606 | NA | NA | NA | NA | 44/606 (7.2%) | NA | NA | NA | NA |
| Dhir[43] | 1141 | NA | NA | NA | NA | 281(25%) | NA | NA | NA | 0/58 (0%) |
| DiMascio[44] | 41 | NA | NA | NA | 1/41 (2.4%) | 14/32 (43.8%) | NA | 1/10 (10%) | NA | 1/41 (2.4%) |
| Diriba[8] | 1271 | NA | NA | NA | 1/72 (1.4%) | 369/686 (53.8%) | NA | 8/69 (11.6%) | NA | 5/430 (1.2%) |
| Duran[51] | 222 | NA | NA | NA | 1/222 (0.45%) | 19/222 (8.5%) | NA | NA | NA | 1/222 (0.45%) |
| Figueiro-Filho[9] | Depends on each outcome | NA | 28/259 (11%) | NA | NA | 159/764 (21%) | 8/241 (3.3%) | 183/992 (18.45%) | NA | 9/1130 (0.8%) |
| Furlan[10] | NA | NA | NA | NA | NA | NA | NA | NA | NA | 1/189 (0.5%) |
| Gajbhiye[11] | 391 | NA | NA | NA | NA | 98/386 (26%) | NA | 31/391 (8%) | NA | 4/369 (1.6 %) |
| Gao[12] | NA | NA | NA | NA | NA | 26/116 (23%) | NA | NA | NA | 1/147 (0.7%) |
| Han[13] | NA | NA | 12/33(30.65%) | NA | 12/160 (18.76%) | 118/743(25.3%) | NA | 108/341(24.4%) | 7/54 (11.1%) | 5/599 (0.8%) |
| Huntley[14] | NA | NA | NA | NA | 1/203 (0.5%) | 57/284 (20.1%) | NA | 137/211 (64.9%) | NA | 1/313 (0.3%) |
| Juan[15] | 221 | NA | NA | NA | NA | NA | NA | 49/173 (28.3%) | NA | 1/221 (0.5%) |
| Kasraeian[16] | Depends on each outcome | NA | NA | NA | NA | 25/41 (60.9%) | NA | NA | NA | 1/86 (0.2%) |
| Khalil[17] | 598 | NA | 45/598 (7.5%) | NA | NA | 198/598 (33.2%) | NA | 179/598 (30%) | NA | 2/528(0.4%) |
| Matar[20] | 136 | 3127.64 g mean | NA | NA | NA | 31/94 (33%) | NA | 27/42 (63.7%) | NA | 3/94 (3.2%) |
| Mirbeyk[21] | 302 | NA | NA | NA | NA | 65 /302 (21.5%) | NA | 5/302 (1.6%) | NA | 2/302 (0.7%) |
| Muhidin[47] | 89 | 1520 to 3820 g range | 7/89 (7.9%) | 2/89 (2.2%) | NA | NA | NA | NA | NA | 2/89 (2.2%) |
| Mullins[22] | 20 | NA | NA | NA | NA | 8/19 (42%) | NA | 1/20 (5%) | NA | 1/20 (5%) |
| Panahi[56] | ** | NA | NA | NA | NA | NA | NA | NA | NA | 1/10 (10%)* |
| Rodriguez Blanco [48] | 74 | NA | NA | NA | 1/74 (1.3%) | NA | NA | NA | NA | 1/74 (1.3%) |
| Segars[49] | 447 | NA | 37/244(15.2%) | NA | 1/17 (5.9%) | 128/236 (54.2%) | NA | NA | NA | 0/91 (0%) |
| Smith[25] | 37 | 2743 g (mean) | 9/21 (42.9 %) | NA | 0/32 (0%) | 6/13 (68.8%) | NA | 11/of 13 (76.9%) | NA | 1/36 (2.7%) |
| Soheili[26] | NA | NA | 13/63 (21%) | NA | NA | 42/151 (28%) | NA | NA | NA | 2/37 (4%) |
| Teles Abrao Trad[27] | 118 | NA | 14/118 ( 11.8%) | NA | NA | 19/118 (16.1%)******** | NA | 24/118 (20.3%) | NA | 1/118 (0.8%) |
| Thomas [28] | 160 | NA | NA | 3/47 (7%) | NA | 24/118 /20%) | NA | NA | NA | 1/160(1%) |
| Trevisanuto[53] | 44 *** | NA | NA | NA | NA | NA | NA | 6/36 (16.6%) | NA | 0/44 (0%) |
| Tripella[29] | 248 | 2914 grams (mean) | NA | NA | 5/190 | 54/196 (27.5%) | NA | NA | NA | 1/248 (0.4%) |
| Trocado[30] | NA | 2292 grams (mean) | 10 (20%) | NA | NA | 18 (35%) | NA | NA | NA | 1/51 (2%) |
| Turan[31] | 479 | NA | NA | 6/479 (1.25%) | 6/361 (1.66%) | 161/479 (24.8%) | NA | 54/479 (11.3%) | NA | 5/479 (1%) |
| Yang N[54] | 84 | NA | 2/38 (5.3%) | NA | NA | 17/80 (21.3%) | NA | NA | NA | 1/84 (1.2%) |
| Yee[34] | 190 | 2855.9 grams (mean) | NA | 9/64 (17.4%) | NA | 54/190 (28.6%) | NA | NA | NA | 1/103 (0.4%) |
| Yoon[35] | 201 | 1880 g to 4050 g range | 15/96 (15.6%) | 5/60 (8.3%) | NA | 48/185 (25.9%) | NA | NA | NA | 1/177 (0.6%) |
| Zaigham[36] | NA | NA | NA | NA | NA | NA | NA | NA | NA | 1/87 (1%) |

* Zhu et al.

** Involves neonates and children up to 16 years/age (it cannot be determined how many neonates are)

***neonates with SARS-CoV-2 infection

**** <36 weeks

### ***Table 6: Vertical transmission to neonates born to SARS COV2 mothers***

| Authors (2020) | N pregnant women | N neonates | Number of sars-cov-2 cases in neonates | Congenital or perinatal | Breastfeeding or breast milk | Respiratory droplets |
| --- | --- | --- | --- | --- | --- | --- |
| AbdelMassih[1] | 1787 | 1787 | 49 | NA | NA | 49 |
| Abdollahpour[39] | NA | NA | 2 | NA | 0/6 (0%) | NA |
| Ashraf[5] |  | 86 | 4 | 0/6 (0%) Placenta  1/16 (6.25%) Amniotic | NA | 4/86 (4.7%) |
| Banaei[40] | NA | NA | 5 | NA | NA | NA |
| Bwire[57] | NA | 206 | 13/206 (6,3%) | NA | NA | NA |
| Caparros Gonzalez[50] | NA | NA | 0/13 (0%) | NA | NA | NA |
| Centeno-Tablante[58] |  | 889 | 124 | NA | 14/82 (17%) | 124/889 (13.9%) |
| Chi[41] | NA | 91 | 8 | NA | NA | NA |
| Deniz[42] | NA | 606 | 20 | 8/63 (12.7%) Placenta  1/8 (12.5%) Amniotic | 3/6 (50%) | NA |
| De Sousa[6] | NA | 493 | 9 | 0/54 (0%) Placenta | NA | NA |
| Dhir[43] | 43 | 148 | 58 | 4/58 (6.9%) Amniotic | NA | 41/58 (70.7%) |
| Di Mascio[44] | NA | 42 | 0/42 (0%) | NA | NA | NA |
| Diriba[8] | 1271 | 1271 | 0/1271 (0%) | NA | NA | NA |
| Duran[51] | NA | 222 | 13 | NA | NA | NA |
| Figueiro-Filho[9] | NA | 1116 | 18/1116 (1,5%) | NA | NA | NA |
| Furlan[10] | NA | 188 | 4 | 0 | NA | NA |
| Gajbhiye[11] | 387 | 313 | 24 | NA | NA | 24/313 (7.7%) |
| Goh[59] | NA | 330 | 9/33 (27.3%) | NA | NA | NA |
| Gordon[52] | NA | 46 | 10 | 0/1 (0%) | NA | 10/46 (21.7%) |
| Han[13] | NA | 559 | 21 | 1/13 (7.7%) Placenta  0/16 (0%) Cord blood  0/17 (0%) Amniotic fluid | 0/10 (0%) | 21/559 (3.8%) |
| Hasan[60] | NA | NA | 0 | NA | NA | NA |
| Hessami[61] | NA | 310 | 0 | NA | NA | NA |
| Juan[15] | NA | 155 | 3/90 (3.3%) | 1/32 (3.1%) Amniotic  0/34 (0%) Cord blood  1/3 (33%) Placenta | 0/22 (0%) | 3/155 (1.9%) |
| Khalil[17] | NA | 751 | 19 | 3/38 (7.9%) Placenta  1/18 (5.6%) Amniotic  1/16 (6.3%) Cord blood | 2/30 (6,6%) | NA |
| Kotlyar[62] | NA | 936 | 27/843 (3.2%) | 2/26 (7.7%) Placenta  0/51 (0%) Amniotic | 2/47 (4,2%) | 27/843 (3,2%) |
| Martins[45] | 24 | 24 | 0 | 0/22 (0%) | 0/24 (0%) | 0/22 (0%) |
| Matar[20] | 136 | 24 | 2/17 (11.5% [95% CI, .067–.192]; I^2^ = 0). | 0/24 (0%) Amniotic  0/24 (0%) Placenta  0/24 (0%) Cord blood. | NA | NA |
| Melo[46] | NA | 405 | 10 | 0/45 (0%) Amniotic  3/26 (11.5%) Placenta  4/28 (14.3%) Cord blood | 0/44 (0%) | 10/405 (2,5%) |
| Mirbeyk[21] | NA | 219 | 11 | 0/219 (0%) Cord blood  0/219 (0%) Placenta  1/219 (0.4%) (Amniotic | 0 | NA |
| Mullins[22] | NA | 15 | 0 | NA | NA | NA |
| Mustafa[63] | NA | 57 | 3 | 0/11 (0%) Amniotic  0/11 (0%) Cord blood  0/2 (0%) Placenta | 0/11 (0%) | 3/57 (5,3%) |
| Pettirosso[23] | NA | 655 | 19 | 4/19 (21%) Placenta  1/19 (5.3%)Cord blood | 0/45 (0%) | 19/655 (2,9%) |
| Rodrí­guez-Blanco[48] | NA | 66 | 0/66 (0%) | NA | 0/6 (0%) | NA |
| Singh[64] | 62 | NA | NA | 2/27 (7,4%) Placenta. | NA | NA |
| Smith[25] | NA | 37 | 1/37 (2.7%) | 0/9 (0%) Cord blood  0/9 (0%) Amniotic | NA | 1/37 (2.7%) |
| Teles Abrao Trad[27] | NA | 95 | 1/95 (1%) | 0 | 0 | 1/95 (1%) |
| Thomas[28] | NA | 81 | 5 | 9/81 (11,1%) Cord blood  9/81 (11,1%) Amniotic | 16/81 (19,8%) | NA |
| Trippella[29] | NA | 191 | 16 | 0/35 (0%) | 0/25 (0%) | 16/191 (8,4%) |
| Turan[31] | 637 | 479 | 8/400 (2%) | 0/20 (0%) Amniotic  0/19 (0%) Cord blood  1/6 (16,7%) Placenta | 1/19 (5,26%) | 8/405 (2%) |
| Yang N[54] | 114 | NA | 2/74 (2,7%) | 0/19 (0%) | 0/17 (0%) | 2/59 (3,4%) |
| Yang Z[55] | NA | 83 | 3/83 (3,6%) | 0/18 (0%) | 0/14 (0%) | 3/62 (4,8%) |
| Yang Z[33] | 16 | 12 | 1 | NA | 0/16 (0%) | 1/12 (8,3%) |
| Yee[34] | NA | 154 | 5 (1,8%) | NA | NA | NA |
| Yoon[35] | 4 | 4 | 4 | 0/4 (0%) Cord blood  0/3 (0%) Amniotic fluid  0/1 (0%) Placenta | 0/4 (0%) | 4/4 (100%) |
| Zaigham[36] | 108 | 75 | 1/75 (1%) | NA | NA | NA |
| Rahman[24] | NA | NA | Narrative description: cases of vertical COVID-19 transmission are few and maybe incidental, the potential of vertical transmission of COVID19 should not be ruled out. | NA | NA | NA |
