## Supplementary material for "COVID-19 and pregnancy: An umbrella review of clinical presentation, vertical transmission, and maternal and perinatal outcomes": S1 to S10: S9 Policy brief of clinical presentation of pregnant women with COVID-19.docx

| 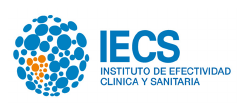 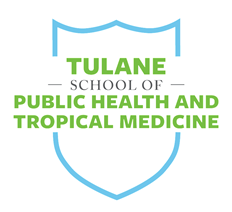 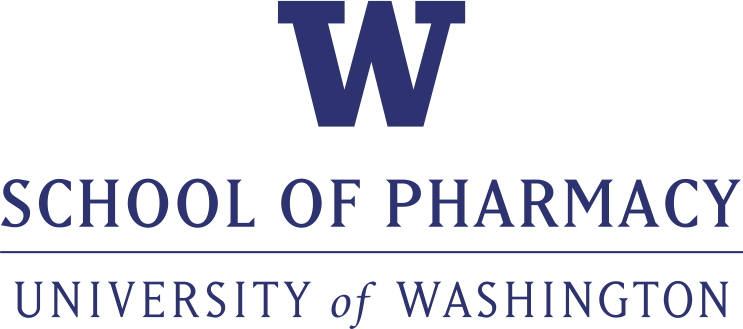 |
| --- |

Who is this summary for?

People deciding how to manage pregnant women with COVID-19

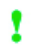
**This summary includes:**

- **Key findings** from research based on a systematic review.
- **Considerations about the relevance of this research** for low- and middle- income countries.

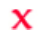
**Not included:**

- Recommendations.
- Additional evidence not included in the systematic review .

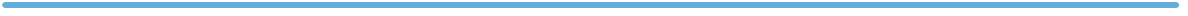

This summary is based on the following systematic review:

Allotey J, Stallings E, Bonet M, et al. Clinical manifestations, risk factors, and maternal and perinatal outcomes of coronavirus disease 2019 in pregnancy: living systematic review and meta-analysis. BMJ (Clinical research ed). 2020;370:m3320.

What is a systematic
review?

A summary of studies addressing a clearly formulated question that uses systematic and explicit methods to identify, select, and critically appraise the relevant research, and to collect and analyse data from the included studies.
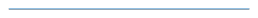

The Institute for Clinical Effectiveness and Health Policy (IECS) is an independent academic institution, affiliated with the Buenos Aires University School of Medicine, devoted to research, education, and technical cooperation in healthcare.

The institution’s activities focus on three complementary fields: research, education and technical cooperation.

October 2020 – Summary of a systematic review

### What is the clinical presentation of pregnant women with COVID-19?

In non-pregnant population with COVID-19 admitted to hospital, the most common symptoms are reported in more than two thirds of the individuals. The clinical patterns in pregnant women with COVID-19 may be different and varies among different settings.

##### Key messages

- A minority of studies were conducted in low- and middle-income countries other than China.

For pregnant women with COVID-19:

- Compared with non-pregnant women of reproductive age with COVID-19, may be less likely to manifest symptoms.
- Fever, cough, dyspnea and ageusia may be the most frequent symptoms.
- Raised C reactive protein level, lymphopenia, raised white cell count and raised procalcitonin level may be the most frequent laboratory findings.
- Ground glass appearance may be the most frequent radiological finding.

#### Background

About the systematic review underlying this summary

**Review objective:** To estimate the effectiveness of professional interventions that, alone or in combination, are effective in antibiotic stewardship for hospital inpatients.

| Types of | What the review authors searched for | What the review authors found |
| --- | --- | --- |
| Study designs & Interventions | Comparative and non-comparative cohort studies with a minimum of 10 participants were included if they reported on the rates and clinical manifestations and relevant outcomes in pregnant and recently pregnant women. | 40 cohort studies reported on clinical manifestations (13,018 pregnant, 85,084 non-pregnant women). |
| Participants | Pregnant and recently pregnant women with suspected or confirmed COVID-19. | Hospitalized and ambulatory patients of any risk. Symptoms (N = 310 to 8328), laboratory findings (N= 251 to 780) and radiological findings (N= 387 to 1968) depending of the type of clinical manifestations. |
| Settings | Hospital and ambulatory settings worldwide. | China (17), USA (7), Spain (4), Italy (3), UK (2), France (2), Netherlands (1), Belgium (1), Brazil (1), Israel (1), Portugal (1). |
| Outcomes | Clinical manifestations. | Proportion of symptoms, laboratory findings and radiological findings. |
| **Date of most recent search:** June 2020 | | |
| **Overall confidence:** Low* **Risk of bias:** Low^#^ | | |

Allotey J, Stallings E, Bonet M, et al. Clinical manifestations, risk factors, and maternal and perinatal outcomes of coronavirus disease 2019 in pregnancy: living systematic review and meta-analysis. BMJ (Clinical research ed). 2020;370:m3320. PMID: [32873575](https://pubmed.ncbi.nlm.nih.gov/32873575/)

*AMSTAR-2: Authors do not provide a list of excluded studies and justify the exclusions nor the sources of funding for the included studies

^#^ROBIS: All items of low risk of bias

How this summary was prepared

After searching widely for systematic reviews that can help inform decisions about COVID-19 & Pregnancy, we have selected the most relevant, highest quality, and most comprehensive one. We used the AMSTAR-2 tool to assess the overall confidence in the results of the review.

The summary formatis based on the [support summaries](https://supportsummaries.epistemonikos.org/home/SearchForm?Search=childhood&locale=en_US&action_results=Search).

A lack of evidence does not mean a lack of effects. It means the effects are uncertain. When there is a lack of evidence, consideration should be given to monitoring and evaluating the effects of the intervention, if it is used.

Testing for SARS-CoV-2 in non-pregnant women is based on symptoms or contact history, but testing in pregnant women is frequently done when they are in hospital for other reasons. However, the particular pattern of COVID-19 related clinical manifestations in pregnant women could be important for diagnosis and management considerations.

#### Summary of findings

40 studies involving 13,018 were included. For symptoms 3 to 29 observational studies and 310 to 8328 participants; for laboratory findings 5 to 15 observational studies and 251 to 780 participants; for radiological findings 10 to 20 observational studies and 387 to 1960 participants.

About the certainty of the evidence (GRADE)

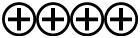

**High:** It is very likely that the effect will be close to what was found in the research.

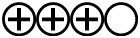

**Moderate:** It is likely that the effect will be close to what was found in the research, but there is a possibility that it will be substantially different.

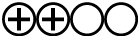

**Low:** It is likely that the effect will be substantially different from what was found in the research, but the research provides an indication of what might be expected.

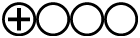

**Very low:** The anticipated effect is very uncertain and the research does not provide a reliable indication of what might be expected.

For pregnant women with COVID-19:

- Compared with non-pregnant women of reproductive age with COVID-19, may be less likely to manifest symptoms (Low certainty of evidence).
- Fever, cough, dyspnea and ageusia may be the most frequent symptoms (Low certainty of evidence).
- Raised C reactive protein level, lymphopenia, raised white cell count and raised procalcitonin level may be the most frequent laboratory findings (Low certainty of evidence).
- Ground glass appearance may be the most frequent radiological finding (Low certainty of evidence).

| **What is the clinical presentation of pregnant women with COVID-19?** | | | |
| --- | --- | --- | --- |
| **Patient or population**: pregnant women with COVID-19  **Setting**: Hospital and ambulatory settings | | | |
| **Outcomes** | **Impact**  **Point estimate (95% confidence interval)** | **№ of participants  (studies)** | **Certainty of the evidence (GRADE)** |
| **Symptoms** Proportion (%); Odds Ratio (Pregnant vs. non-pregnant) | Fever 40 % (31 to 49); OR 0.43 (0.22 to 0.85)  Cough 39 % (31 to 47); OR 0.67 (0.37 to 1.23)  Dyspnea 19 % (13 to 26); OR 0.82 (0.47 to 1.43)  Ageusia 15 % (0 to 41); -  Myalgia 10 % (5 to 17); OR 0.48 (0.45 to 0.51)  Diarrhea 7 % (5 to 9); -  Any symptom - ; OR 0.33 (0.08 to 1.41) | 310 to 8328  (3 to 29 observational studies) | 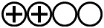  Low ^a,b^ |
| **Laboratory findings**  Proportion (%) | Raised C reactive protein level 49 % (36 to 63)  Lymphopenia 35 % (26 to 45)  Raised white cell count 27 % (9 to 51)  Raised procalcitonin level 21 % (0 to 59)  Abnormal liver function test results 11 % (5 to 18)  Thrombocytopenia 8 % (2 to 18) | 251 to 780  (5 to 15 observational studies) | 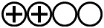  Low ^a,c^ |
| **Radiological findings**  Proportion (%) | Ground glass appearance 69 % (41 to 91)  Any abnormality on computed tomography 0 % (0 to 1) | 387 to 1960  (10 to 20 observational studies) | 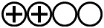  Low ^a,d^ |
| **GRADE Working Group grades of evidence** **High certainty:** We are very confident that the true proportions or association effect lies close to that of the estimate **Moderate certainty:** We are moderately confident in the proportions or association effect estimate: The true proportions or association effect is likely to be close to the estimate, but there is a possibility that it is substantially different **Low certainty:** Our confidence in the proportions or association effect estimate is limited: The true proportions or association effect may be substantially different from the estimate **Very low certainty:** We have very little confidence in the proportions or association effect estimate: The true proportions or association effect is likely to be substantially different from the estimate  In the field of prognosis, a body of observational evidence begins as high-quality evidence, and the same concept applies to prevalence studies.^1^  1. Iorio A, Spencer FA, Falavigna M, et al. Use of GRADE for assessment of evidence about prognosis: rating confidence in estimates of event rates in broad categories of patients. BMJ. 2015;350:h870. PMID: [25775931](https://pubmed.ncbi.nlm.nih.gov/25775931/) DOI: [10.1136/bmj.h870](https://www.bmj.com/content/350/bmj.h870) | | | |

**a**. Low to moderate risk of bias; **b**. I^2^ from 66 to 98% (lower heterogeneity of point estimates); c. I^2^ 74 to 97% (lower heterogeneity of point estimates); d. I^2^ 97 to 98% (lower heterogeneity of point estimates)

#### Relevance of the review for low and middle-income countries

| 🡺 Findings | ▷ Interpretation* |
| --- | --- |
| APPLICABILITY | |
| 🡺 Only 1 out of 40 studies included in the systematic review, other than China, were conducted in low and middle-income countries. | ▷ The estimations of clinical manifestations may vary in different countries. |
| EQUITY | |
| **🡺 There was no information in the included studies regarding the different clinical manifestations on resource-disadvantaged populations.** | ▷ The estimations of clinical manifestations may vary in different settings, affecting the diagnosis and management. |
| ECONOMIC CONSIDERATIONS | |
| **🡺 The systematic review did not address economic considerations.** | ▷ Local costings laboratory and radiological tests should be undertaken, particularly in settings differing from the original investigations. |
| MONITORING & EVALUATION | |
| **🡺 There is low-certainty evidence the proportion of symptoms, laboratory findings and radiological findings regarding the first pandemic wave.**  **🡺 There is no evidence regarding the second pandemic wave.** | ▷ Larger and more rigorous studies should provide more evidence from low and middle-income countries and from resource-disadvantaged populations.  ▷ New evidence regarding the second pandemic wave will be required. |

*Judgments made by the authors of this summary, not necessarily those of the review authors, based on the findings of the review and consultation with researchers and policymakers in low-income countries. For additional details about how these judgments were made see:
[www.supportsummaries.org/methods](http://www.supportsummaries.org/methods)

#### Additional information

About certainty of the evidence (GRADE)

The “certainty of the evidence” is an assessment of how good an indication the research provides of the likely effect; i.e. the likelihood that the effect will be substantially different from what the research found. By “substantially different” we mean a large enough difference that it might affect a decision. These judgements are made using the GRADE system, and are provided for each outcome. The judgements are based on the study design (randomised trials versus observational studies), factors that reduce the certainty (risk of bias, inconsistency, indirectness, imprecision, and publication bias) and factors that increase the certainty (a large effect, a dose response relationship, and plausible confounding). For each outcome, the certainty of the evidence is rated as high, moderate, low or very low using the definitions on page 3.

**For more information about GRADE:**

[www.gradeworkinggroup.org](http://www.gradeworkinggroup.org)

About applicability

Blah blah genereal text about this. These findings to other lower and middle income countries. Integrated Management of Childhood Illness comprises.

About equity

The quality of the evidence indicated in the table

About scaling up

The quality of the evidence indicated in the table

**Glossary of terms** used in this report: [www.support.org/explanations.htm](http://www.support.org/explanations.htm)

**Receive e-mail notices of new SUPPORT summaries**: [www.support.org/newsletter.htm](http://www.support.org/newsletter.htm)

###### Related literature

Eight out of 20 systematic reviews assessing clinical manifestations patterns among pregnant women with COVID-19, provided additional results to the summarized systematic review, but all included fewer studies and participants. In general, described mild to moderate manifestations of COVID-19^1,3^, adding fatigue^4^, elevated D-dimer^5^, elevated transaminases^6^ and CT findings^7,8^.

1. Trippella G, Ciarcià M, Ferrari M, et al. COVID-19 in Pregnant Women and Neonates: A Systematic Review of the Literature with Quality Assessment of the Studies. Pathogens (Basel, Switzerland). 2020;9(6):1-29.
2. Turan O, Hakim A, Dashraath P, Jeslyn WJL, Wright A, Abdul Kadir R. Clinical characteristics, prognostic factors, and maternal and neonatal outcomes of SARS-CoV-2 infection among hospitalized pregnant women: a systematic review. International journal of gynaecology and obstetrics: the official organ of the International Federation of Gynaecology and Obstetrics. 2020.
3. Rodríguez-Blanco N, Vegara-Lopez I, Aleo-Giner L, Tuells J. [Scoping review of coronavirus case series (SARS-CoV, MERS-CoV and SARS-CoV-2) and their obstetric and neonatal results]. Revista espanola de quimioterapia : publicacion oficial de la Sociedad Espanola de Quimioterapia. 2020.
4. Khan MA, Khan N, Mustagir G, Rana J, Haque R, Rahman M. COVID-19 infection during pregnancy: A systematic review to summarize possible symptoms, treatments, and pregnancy outcomes. medRxiv. 2020:2020.2003.2031.20049304.
5. Shi L, Wang Y, Yang H, Duan G. Laboratory Abnormalities in Pregnant Women with Novel Coronavirus Disease 2019. American journal of perinatology. 2020;37(10):1070-1073.
6. Khalil A, Kalafat E, Benlioglu C, et al. SARS-CoV-2 infection in pregnancy: A systematic review and meta-analysis of clinical features and pregnancy outcomes. EClinicalMedicine. 2020:100446.
7. Soheili M, Moradi G, Baradaran HR, Soheili M, Moradi Y. Clinical Manifestation and Maternal Complications and Neonatal outcomes in Pregnant Women with COVID 19: An Update a Systematic Review and Meta-analysis. In: Research Square; 2020.
8. Gao YJ, Ye L, Zhang JS, et al. Clinical features and outcomes of pregnant women with COVID-19: a systematic review and meta-analysis. BMC infectious diseases. 2020;20(1):564.

###### This summary was prepared by

Agustín Ciapponi, Ariel Bardach, Agustina Mazzoni, Instituto de Efectividad Clínica y Sanitaria (IECS-CONICET), Buenos Aires, Argentina

###### Conflict of interest

None declared.

###### Acknowledgements

This summary has been peer reviewed by Sarah Mathews, USA.

###### This review should be cited as

Allotey J, Stallings E, Bonet M, et al. Clinical manifestations, risk factors, and maternal and perinatal outcomes of coronavirus disease 2019 in pregnancy: living systematic review and meta-analysis. BMJ (Clinical research ed). 2020;370:m3320.

###### The summary should be cited as

###### Ciapponi A, Mazzoni A, Bardach A. What is the clinical presentation of pregnant women with COVID-19? Summary of a systematic review. October 2020. [www.iecs.org.ar](http://www.iecs.org.ar)

###### [Financial support](https://www.gatesfoundation.org/)

[Grant of Bill & Melinda Gates Foundation](https://www.gatesfoundation.org/)

**Keywords**

All Summaries:

evidence-informed health policy, evidence-based, systematic review, COVID-19, pregnancy, low and middle-income countries, maternal health clinical presentation, symptoms.
