## Supplementary material for "COVID-19 and pregnancy: An umbrella review of clinical presentation, vertical transmission, and maternal and perinatal outcomes": S1 to S10: S10 Policy brief of transmission of SARS-CoV-2 through breastfeeding.docx

| 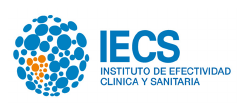 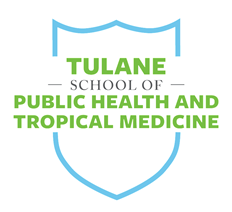 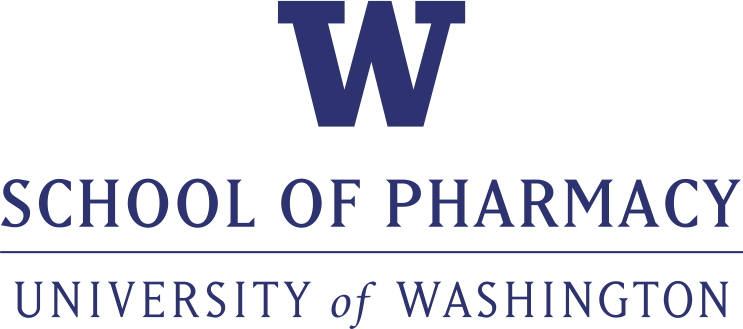 |
| --- |

Who is this summary for?

Those making decisions about breastfeeding recommendations for women with COVID-19

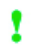
**This summary includes:**

- **Key findings** from research based on a systematic review
- **Considerations about the relevance of this research** for low- and middle- income countries

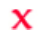
**Not included:**

- Recommendations
- Additional evidence not included in this systematic review

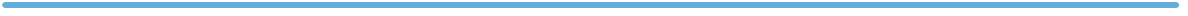

This summary is based on the following systematic review:

Centeno-Tablante E, Medina-Rivera M, Finkelstein JL, et al. Transmission of SARS-CoV-2 through breast milk and breastfeeding: a living systematic review. Ann N Y Acad Sci. 2020.

What is a systematic
review?

A summary of studies addressing a clearly formulated question that uses systematic and explicit methods to identify, select, and critically appraise the relevant research. The objective is to collect and analyse data from the included studies.
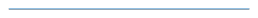

The Institute for Clinical Effectiveness and Health Policy (IECS) is an independent academic institution, affiliated with the Buenos Aires University School of Medicine, devoted to research, education, and technical cooperation in healthcare.

The institution’s activities focus on three complementary fields: research, education and technical cooperation.

October 2020 – Summary of a systematic review

### Do mothers transmit SARS-CoV-2 infection to their offspring through breastfeeding?

**Summary includes:**

**- Summary of research findings**, based on one or more systematic reviews of research on this topic

**- Relevance** for low and middle income countries

**Doesn’t include:**

- Recommendations

- Cost assessments

- Results from qualitative stuides

- Examples or detailed descriptions of implementation

Immediate breastfeeding provides many benefits to infants, including reducing the risk of neonatal mortality, especially in low- and middle-income countries. Guidance on breastfeeding for mothers with COVID-19 could have a huge public health impact.

##### Key messages

- There have been few studies conducted in a low- and middle-income countries other than China regarding the risk of breastfeeding among mothers with COVID-19.
- SARS‐CoV‐2 transmission via breast milk is very uncertain, but published results suggested that the risk, at most, would be low.
- Transmission via breastfeeding through other related bodily fluids, (ie. droplet transmission or airborne transmission due to close contact with the infant or young child) could pose a risk to the infant. However, the evidence is very uncertain.

#### Background

About the systematic review underlying this summary

**Review objective:** To assess available evidence related to the possible transmission of SARS‐CoV‐2 through breast milk and breastfeeding (i.e., directly through breast milk or through close-contact with related bodily fluids, such as blood, sweat, and respiratory droplets).

|  | What the review authors searched for | What the review authors found |
| --- | --- | --- |
| Study designs & Interventions | Any study design, including case reports, case series, report of family clusters and cohort studies. | 37 studies analyzed breast milk samples: 28 case reports and 9 retrospective case series.^#^ |
| Participants | Pregnant or lactating women with suspected, probable, or confirmed SARS‐CoV‐2 infection as well as their infants or young children (0–24 months of age) consuming breast milk directly from the breast or extracted breast milk. | 77 infants whose mothers were diagnosed with COVID‐19 and were able to provide a breast milk specimen for analysis. |
| Settings | Hospital and ambulatory settings worldwide. | China (21), Italy (6), Germany (2), Turkey (2), and 1 each from Australia, Belgium, Canada, Jordan, Singapore, and the Republic of Korea. |
| Outcomes | Infant with suspected, probable, or confirmed SARS‐CoV‐2 infection within 30 days of breastfeeding or receiving expressed breast milk from a woman with a suspected, probable, or confirmed SARS‐CoV‐2 infection. | 17 out of the 19 positive children with described infant feeding practices (10 cases were reported to be breastfed, 4 mix‐fed, 2 cases received breast milk substitute). |
| **Date of most recent search:** July 07, 2020 | | |
| **Overall confidence:** Critically Low* **Risk of bias:** Unclear^#^ | | |

Centeno-Tablante E, Medina-Rivera M, Finkelstein JL, et al. Transmission of SARS-CoV-2 through breast milk and breastfeeding: a living systematic review. Ann N Y Acad Sci. 2020.. PMID: [32860259](https://pubmed.ncbi.nlm.nih.gov/32860259/)

^#^ We did not include the 303 reports without breast milk samples tested since it is not possible to ascertain if there is an increased risk of viral infection among breastfeeding children via breast milk.

* Authors do not provide a list of excluded studies and justify the exclusions, they used GRADE but there is no statement about the tool to classify the risk of bias domain of each study, and they do not provide the sources of funding for the included studies.

^#^ROBIS: All items of low risk of bias but unclear for synthesis and findings domain.

How this summary was prepared

After searching widely for systematic reviews that can help inform decisions about COVID-19 & pregnancy, we have selected the most relevant, comprehensive publications of the highest quality. We used the AMSTAR-2 tool to assess the overall confidence in the results of the review.

The summary format is based on the [support summaries](https://supportsummaries.epistemonikos.org/home/SearchForm?Search=childhood&locale=en_US&action_results=Search).

A lack of evidence does not mean a lack of effect, but rather that the potential effects are uncertain. When there is a lack of evidence, consideration should be given to monitoring and evaluating the effects of interventions, if used.

On May 27, 2020, the World Health Organization (WHO) recommended exclusive breastfeeding for mothers with suspected or confirmed COVID-19 infection for at least the first 6 months and breastfeeding alongside complementary foods until 2 years of age, taking necessary precautioins. This recommendation was based on the health benefits associated with breastfeeding for both the mother and the child and the observed relatively mild or asymptomatic illness of COVID-19 in infants. Evidence regarding safe infant feeding practice are very important for families and health personnel in the current pandemic context.

#### Summary of findings

Among the included studies with laboratory‐confirmed breast milk samples from 77 mothers, the **breastfed group** composed of **8** COVID‐19 neonates (first 4 weeks after birth), 15 neonates without COVID‐19, **2** confirmed cases infants (3 and 6 months old); the **mix‐fed group** had **2** positive neonate, 2 negative neonates and **3** breastfed infants (2, 8 and 14 months old); in the **formula group** (including infants separated from their mothers after birth) there were **2** positive infected neonates, 16 negative neonates, and no infected infants. Infant feeding practices were **not reported** for **2** positive neonates nor for 25 neonates who were negative for COVID‐19.

About the certainty of the evidence (GRADE)

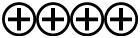

**High:** It is very likely that the effect will be close to what was found in the research.

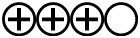

**Moderate:** It is likely that the effect will be close to what was found in the research, but there is a possibility that it will be substantially different.

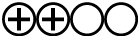

**Low:** It is likely that the effect will be substantially different from what was found in the research, but the research provides an indication of what might be expected.

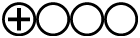

**Very low:** The anticipated effect is very uncertain and the research does not provide a reliable indication of what might be expected.

In all cases of children with COVID‐19, transmission could have occurred through alternatic routes such as blood, sweat, and respiratory droplets, skin‐to‐skin contact, airborne transmission or close contacts with infected family members. In fact, several confirmed COVID‐19 infants by RT‐PCR tests received breast milk negative for SARS‐CoV‐2 from COVID‐19 mothers.

- SARS‐CoV‐2 transmission via breast milk is very uncertain and the risk of transmission via this route is estimated to be, at most, low (Very low certainty of evidence).
- Transmission via breastfeeding through other related bodily fluids, (ie. droplet transmission or airborne transmission due to close contact with the infant or young child) could pose a risk to the infant. However, the evidence is very uncertain (Very low certainty of evidence).

| **Do mothers transmit SARS-CoV-2 infection to children through breastfeeding?** | | | |
| --- | --- | --- | --- |
| **Patient or population**: Mothers with COVID-19 able to breastfeed  **Setting**: Hospital and ambulatory settings | | | |
| **Outcomes** | **Impact**  **Point estimate (95% confidence interval)** | **№ of participants  (studies)** | **Certainty of the evidence (GRADE)** |
| **Proportion of children with COVID‐19 from mothers with COVID-19 who are able to breastfeed** (detected by RT‐PCR analysis) | 19*/77 (25%) children whose mothers were diagnosed with COVID‐19 were confirmed COVID‐19 cases (14 neonates and 5 older infants).  4/6 (67%) children exposed to positive breast milk samples were confirmed COVID‐19 cases (3 neonates and 1 infant).  The proportion of children COVID‐19 by feeding practice was:  10/23 (43%) of children that received breastfeeding;  2/18 (11%) of neonates that received breast milk substitute;  5/7 (71%) of children that received mix-feeding;  2/27 (7%) of children without information about their feeding practice.  SARS‐CoV‐2 transmission via breast milk was not confirmed in any case. | 77 children  (37 observational studies) | 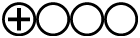  Very low ^a,b,c^ |
| **Breast milk analysis for SARS‐CoV‐2 RNA positivity** | 9**/**82 (11%) analyzed breast milk samples from mothers with COVID‐19 were positive for antibodies or SARS‐CoV‐2 RNA (9/68 tested for SARS‐CoV‐2 RNA).  The viral culture of 2 breast milk samples with viral RNA was negative. | 82 breast milk samples  (37 observational studies) | 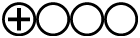  Very low ^a,b,c^ |
| **GRADE Working Group grades of evidence** **High certainty:** We are very confident that the true proportions or association effect lies close to that of the estimate **Moderate certainty:** We are moderately confident in the proportions or association effect estimate: The true proportions or association effect is likely to be close to the estimate, but there is a possibility that it is substantially different **Low certainty:** Our confidence in the proportions or association effect estimate is limited: The true proportions or association effect may be substantially different from the estimate **Very low certainty:** We have very little confidence in the proportions or association effect estimate: The true proportions or association effect is likely to be substantially different from the estimate  In the field of prognosis, a body of observational evidence begins as high-quality evidence, and the same concept applies to prevalence studies.^1^  1. Iorio A, Spencer FA, Falavigna M, et al. Use of GRADE for assessment of evidence about prognosis: rating confidence in estimates of event rates in broad categories of patients. BMJ. 2015;350:h870. PMID: [25775931](https://pubmed.ncbi.nlm.nih.gov/25775931/) DOI: [10.1136/bmj.h870](https://www.bmj.com/content/350/bmj.h870) | | | |

**a**. All studies had a high risk of bias due to lack of a control group, short follow‐up time, and lack of control for other possible confounders, Additionally, the sample could also be biased by only testing infants who seek care and present with severe symptoms.; **b**. Imprecision due to few cases; c. high risk of publication bias is plausible given that COVID‐19–positive cases of infants and breast milk are most likely to be reported and published.

* One newborn was diagnosed by the detection of anti‐SARS‐CoV–2–specific antibodies in serum

#### Relevance of the review for low- and middle-income countries

| 🡺 Findings | ▷ Interpretation* |
| --- | --- |
| APPLICABILITY | |
| 🡺 Only 2 out of 37 studies included in the systematic review, other than China, were conducted in low- and middle-income countries. | ▷ Mother‐to‐child transmission may vary in different countries depending the hospital and home possibilities to prevent COVID-19. |
| EQUITY | |
| **🡺 There was no information in the included studies regarding mother‐to‐child transmission in resource-disadvantaged populations.**  **🡺 Most studies tested infants who seek care and presented with severe symptoms.** | ▷ Mother‐to‐child transmission may be higher in low resource settings.  ▷ A reduction of breastfeeding could have a greater impact in resource-disadvantaged populations than other groups due to lack of breastfeeding benefits.  ▷ Diagnosis and management could be more difficult in resource-disadvantaged populations. |
| ECONOMIC CONSIDERATIONS | |
| **🡺 This systematic review did not address economic considerations.** | ▷ Feeding practices can have a big impact on families’ economic well-being and on public health. |
| MONITORING & EVALUATION | |
| **🡺 The evidence about the magnitude and mechanisms of potential SARS‐CoV‐2 breastfeeding transmission is of very low certainty.**  **🡺 There is no evidence regarding the second wave of coronavirus infections.** | ▷ Larger and more rigorous studies should provide more evidence from low and middle-income countries and from resource-disadvantaged populations.  ▷ Studies should confirm and detail clinical characteristics and timing of maternal and infant infection; analyze and report on the presence of virus and viral infectious particles in breast milk, ideally with serial samples; record data on infant feeding practices and any contact precautions taken, such as wearing a mask and isolation measures. |

*Judgements made by the authors of this summary, not necessarily those of the review authors, based on the findings of the review and consultation with researchers and policymakers in low-income countries. For additional details about how these judgements were made see:
[www.supportsummaries.org/methods](http://www.supportsummaries.org/methods)

#### Additional information

About certainty of the evidence (GRADE)

The “certainty of the evidence” is an assessment of how good an indication the research provides of the likely effect; i.e. the likelihood that the effect will be substantially different from what the research found. By “substantially different” we mean a large enough difference that it might affect a decision. These judgements are made using the GRADE system, and are provided for each outcome. The judgements are based on the study design (randomised trials versus observational studies), factors that reduce the certainty (risk of bias, inconsistency, indirectness, imprecision, and publication bias) and factors that increase the certainty (a large effect, a dose response relationship, and plausible confounding). For each outcome, the certainty of the evidence is rated as high, moderate, low or very low using the definitions on page 3.

**For more information about GRADE:**

[www.gradeworkinggroup.org](http://www.gradeworkinggroup.org)

About applicability

Blah blah genereal text about this. These findings to other lower and middle income countries. Integrated Management of Childhood Illness comprises.

About equity

The quality of the evidence indicated in the table

About scaling up

The quality of the evidence indicated in the table

**Glossary of terms** used in this report: [www.support.org/explanations.htm](http://www.support.org/explanations.htm)

**Receive e-mail notices of new SUPPORT summaries**: [www.support.org/newsletter.htm](http://www.support.org/newsletter.htm)

###### Related literature

Four systematic reviews assessing breastfeeding mother‐to‐child transmission among women with COVID-19 infection, provided few additional findings to supplement the summarized systematic review. However, all included fewer studies and participants. One review analyzed the presence of SARS-CoV-2 RNA in the breast milk of 24 pregnant women with COVID-19 during the third trimester of pregnancy without founding evidence of SARS-CoV-2^1,^. Two systematic reviews including five studies each reported negative results for SARS-CoV-2 in breastmilk^2,3^. Another review included 201 infants neonates born to mothers with COVID-19 and four of them were reported to have laboratory-confirmed SARS-CoV-2 infection within 48 hours after birth. However, the RT‐PCR tests of the breast milk, placenta, amniotic fluids, and cord blood and maternal vaginal secretions were all negative for SARS-CoV-2 in the reported cases^4^. The Union of European Neonatal & Perinatal Societies (UENPS) after reviewing the evidence and recommendations of other organizations recommended breastfeeding for asymptomatic or paucisymptomatic mothers COVID-19 positive at delivery under strict measures of infection control. When a mother with COVID-19 is too sick to care for the newborn, the neonate should be managed separately and fed fresh expressed breast milk, with no need to pasteurize it^5^.

###### This summary was prepared by

Agustín Ciapponi, Agustina Mazzoni, Ariel Bardach. Instituto de Efectividad Clínica y Sanitaria (IECS-CONICET), Buenos Aires, Argentina

###### Conflict of interest

None declared.

###### Acknowledgements

This summary has been peer reviewed by Sarah Mathews, USA.

###### This review should be cited as

Centeno-Tablante E, Medina-Rivera M, Finkelstein JL, et al. Transmission of SARS-CoV-2 through breast milk and breastfeeding: a living systematic review. Ann N Y Acad Sci. 2020.

###### The summary should be cited as

###### Ciapponi A, Mazzoni A, Bardach A. Do mothers transmit SARS-CoV-2 infection to their offspring through breastfeeding? Summary of a systematic review. October 2020. <http://safeinpregnancy.org/>

###### [Financial support](https://www.gatesfoundation.org/)

[Grant of Bill & Melinda Gates Foundation](https://www.gatesfoundation.org/)

**Keywords**

All Summaries:

evidence-informed health policy, evidence-based, systematic review, COVID-19, SARS‐CoV‐2, breastfeeding, human milk, breast milk, vertical transmission, perinatal transmission, mother‐to‐child transmission, low and middle-income countries, child health.
